# Using genomic epidemiology of SARS-CoV-2 to support contact tracing and public health surveillance in rural Humboldt County, California

**DOI:** 10.1101/2021.09.21.21258385

**Authors:** Gunnar Stoddard, Allison Black, Patrick Ayscue, Dan Lu, Jack Kamm, Karan Bhatt, Lienna Chan, Amy L Kistler, Joshua Batson, Angela Detweiler, Michelle Tan, Norma Neff, Joseph L DeRisi, Jeremy Corrigan

**Author notes:** These authors contributed equally to this article (Gunnar Stoddard, Allison Black).

## Abstract

During the COVID-19 pandemic within the United States, much of the responsibility for diagnostic testing and epidemiologic response has relied on the action of county-level departments of public health. Here we describe the integration of genomic surveillance into epidemiologic response within Humboldt County, a rural county in northwest California. Through a collaborative effort, 853 whole SARS-CoV-2 genomes were generated, representing ∼58% of the 1,449 SARS-CoV-2-positive cases detected in Humboldt County as of mid-March 2021. Phylogenetic analysis of these data was used to develop a comprehensive understanding of SARS-CoV-2 introductions to the county and to support contact tracing and epidemiologic investigations of all large outbreaks in the county. In the case of an outbreak on a commercial farm, viral genomic data were used to validate reported epidemiologic links and link additional cases within the community who did not report a farm exposure to the outbreak. During a separate outbreak within a skilled nursing facility, genomic surveillance data were used to rule out the putative index case, detect the emergence of an independent Spike:N501Y substitution, and verify that the outbreak had been brought under control. These use cases demonstrate how developing genomic surveillance capacity within local public health departments can support timely and responsive deployment of genomic epidemiology for surveillance and outbreak response based on local needs and priorities.

## INTRODUCTION

In early December 2019 a novel human coronavirus, now known as SARS-CoV-2, emerged in Wuhan, China. During January and February 2020, travel-associated cases of COVID-19, the disease caused by SARS-CoV-2, surfaced within the United States. By the end of February 2020 the United States reported instances of community transmission (Ghinai et al. 2020). Over the following year, the scope of the resulting pandemic has become clear. There have been greater than 33 million reported cases nationally, and 3.7 million just in California as of this writing (May 2021).

Within the United States, frontline public health activities typically fall under the jurisdictional authority of states, counties, or smaller administrative units. County-level public health departments are routinely responsible for performing diagnostic testing, mounting epidemiologic responses, and reporting notifiable illnesses or conditions to higher level jurisdictional authorities. Thus, much of the responsibility for detecting and preventing COVID-19 transmission has relied on the action of county-level departments of public health. Notably, despite a remit to conduct similar public health activities, the capacities and resources available to departments of public health varies greatly across the United States and has inevitably resulted in an uneven response to the pandemic.

When compared to other counties in California, the unique features of Humboldt County present particular challenges with respect to pandemic response, especially for COVID-19. Humboldt County is a rural county in northwest California encompassing 2.3 million acres, 80 percent of which is forestland, protected redwoods and recreation areas, serviced by a single small regional airport. The county is a healthcare and behavioral healthcare provider shortage area, meaning that many of the county’s 135,727 residents must travel long distances for access to healthcare services, and the largest population center (Eureka-Arcata) may be isolated due to landslides or other natural disasters. Many Humboldt residents are uninsured and more than one in five individuals live at or below the federal poverty line. The largest minority populations are Native American and Latino, both at high risk for serious COVID-19 disease, and the area experiences notable health disparities and overall poorer health outcomes when compared to state and national data (Humboldt County Department of Health & Human Services, Public Health 2018). Furthermore, pandemic preparedness has been hampered by a dearth of laboratory facilities in the region. The Humboldt County Public Health Laboratory (HCPHL) was the only local testing laboratory for SARS-CoV-2 early in the pandemic, with local hospitals and clinics adding testing options subsequently.

Despite these challenges, Humboldt County has used genomic sequencing of SARS-CoV-2 as a key component in the arsenal of epidemiological tools used to monitor, track, and control spread of the virus. Epidemiological analyses of viral sequence data have played critical roles in our understanding of SARS-CoV-2 epidemiology, and have been used to detect cryptic transmission of SARS-CoV-2 (Bedford et al. 2020; Alteri et al. 2021), identify multiple, independent introductions of SARS-CoV-2 (Laiton-Donato et al. 2020; Alteri et al. 2021; Badaoui et al. 2021), characterize SARS-CoV-2 transmission patterns domestically within the United States (Fauver et al. 2020), and calculate the increased transmissibility of newly detected variants (Davies et al. 2021), including here in California (Peng et al. 2021). Despite their value, the technical complexity of genomic surveillance systems has often limited their accessibility outside of higher resource urban settings, including at the county-level within the United States. In this paper we describe how, within a rural county locale, we improved diagnostic testing capacity and implemented dense genomic surveillance of SARS-CoV-2. We then discuss how findings from genomic surveillance data supported public health surveillance and guided epidemiologic response efforts in Humboldt County, with specific attention on two outbreaks, one in a farm setting, and the other in the context of a skilled nursing facility. Humboldt County’s pandemic response can offer a roadmap for other local jurisdictions outside urban centers evaluating how to incorporate genomic epidemiology into their response efforts.

## METHODS

### Ethics statement

This work was approved by the University of California San Francisco Human Research Protection Program Institutional Review Board (IRB# 21-34522, Reference Number 319235). This same IRB also waived consent for specimen collection because collection occurred as part of public health response. All samples submitted to the Chan Zuckerberg Biohub for sequencing were de-identified. All genomic analysis was performed on de-identified genome sequences and sample metadata. All work was carried out in accordance with relevant regulations and guidelines.

### Collection of SARS-CoV-2 diagnostic specimens

Samples were collected from a variety of submitters throughout Humboldt County including local hospitals, clinics, skilled nursing facilities (SNFs), assisted living facilities (ALFs), other congregate living facilities such as behavioral health centers, the county jail, local college dormitories and local health centers. Samples were typically collected upon clinical suspicion of SARS-CoV-2 infection or as part of case investigations conducted by the Humboldt County Department of Health and Human Services - Public Health (DHHS-PH), which performed targeted prospective surveillance in populations of concern such as SNFs and ALFs. While some SNFs were subject to routine surveillance testing to identify possible outbreaks early on, most samples were collected from symptomatic individuals and thus represent a skewed sample of all infections that likely occurred within Humboldt County.

Throughout the pandemic the sample type evolved. Initially, samples were collected using dual or combined nasopharyngeal/oropharyngeal swabs. More recently, nasopharyngeal swabs and observed self-collected nasal swabs were the primary sample types that were received and tested. Additionally, swabs in both viral transport media and saline transport media were validated for testing. Samples were collected and transported to the lab as soon as possible and held at 4 degrees Celsius for up to 72 hrs, or frozen for up to 7 days if sample processing was delayed.

### Logistics of SARS-CoV-2 testing in Humboldt County

Early in the pandemic the majority of specimens collected by health care providers within Humboldt County requiring SARS-CoV-2 testing were handled by the HCPHL. A prioritization system was established to ensure that symptomatic samples and high-risk populations were tested locally, which improved result turnaround times. Low priority samples, such as those collected to provide travel clearance, pre-operative screening, and general population surveillance, were deferred to commercial laboratories such as Quest and LabCorp. SARS-CoV-2 testing of hospital in-patients was mostly conducted by the hospital. Additionally, some specimens were collected and tested by OptumServe through a partnership with the Californian government to increase testing capacity.

The OptumServe testing site served as a community surveillance site and began operating in April 2020. The site was initially a walk-in sample collection service that operated 5 days per week, collecting approximately 120 samples per day. Optum sample collection capacity increased over time, expanding to two collection teams which increased capacity up to 319 samples per day. Additionally, an Optum mobile team provided capacity to collect up to 561 samples per week. All samples collected by Optum were tested at the California Department of Public Health’s Valencia Branch Laboratory. None of these samples were sequenced as part of this project. Finally, Humboldt County, Del Norte County and United Indian Health Services formed a regional COVID testing task force to coordinate testing strategy and resources for the region. Analysis by the task force led to the establishment of the North Coast Testing Partnership (NCTP) which began performing diagnostic testing in December 2020. The NCTP lab tested between 100-200 samples 5 days a week.

### Viral RNA extraction

Throughout the pandemic the HCPHL used a variety of extraction platforms. RNA extractions were initially performed manually using the QIAamp Viral RNA mini kit (Qiagen) following manufacturer instructions. To improve throughput, automated RNA extraction was implemented on the MagNAPure Compact (Roche), eluting the extracted sample in 100 µls of elution buffer. Additionally, some extractions were either performed on the Qiagen EZ1 Advanced, eluting samples in 120 µls of elution buffer, or on the KingFisher™ Flex Purification System and the MagMAX™ Viral/Pathogen II Nucleic Acid Isolation Kit (ThermoFisher), which eluted the sample in 100 µls of elution buffer.

### SARS-CoV-2 diagnostic testing

At the beginning of the pandemic, HCPHL used the CDC 2019-Novel Coronavirus (2019-nCoV) Real-Time RT-PCR Diagnostic Panel. This assay is a singleplex assay which only allows processing of 10 samples at a time. In August of 2020 we began using the TaqPath COVID-19 Combo Kit (ThermoFisher), which allowed multiplexing of 93 samples per run. In preparation for Influenza season, HCPHL transitioned to the CDC Influenza SARS-CoV-2 (Flu SC2) Multiplex Assay that identifies SARS-CoV-2, Influenza A, and Influenza B simultaneously and tests 93 samples per run. Recent diagnostic testing primarily used the CDC Flu SC2 Multiplex Assay. To accommodate rapid turnaround of high-priority specimens, the HCPHL also validated the Xpert Xpress SARS-CoV-2 assay using the GeneXpert testing system (CEPHEID). The lab upgraded the GeneXpert testing system from 4 modules to 16 modules to increase capacity and reduce turn-around time. Additionally, the Xpert SARS-CoV-2/Flu/RSV 4-plex assay (CEPHEID) was validated and used during the influenza season. All PCR assays were used according to manufacturer instructions and instructions for use (IFU) without deviation.

### Selection of samples for sequencing

Convenience samples of diagnostic specimens were sent periodically for whole genome sequencing at the Chan Zuckerberg Biohub in San Francisco. Viral load was used as the primary criteria for choosing samples to sequence given that samples with higher viral loads are more likely to yield high quality whole genome sequences. All samples with RT-PCR cycle threshold (Ct) values of less than 31 were selected. Additional samples of epidemiologic interest, such as specimens associated with specific case investigations, were also included. Eluted RNA was aliquoted into 96-well plates and sent to the Biohub in batches ranging from 40 to 96 samples. Metadata were compiled by communicable disease investigators to assist in post-sequencing analysis and case investigations.

### Sequencing of SARS-CoV-2 whole genomes

Extracted total nucleic acid was diluted based on average SARS-CoV-2 N and E gene cycle threshold (Ct) values; samples with a Ct range 12-15 were diluted 1:100, 15-18 1:10 and >18 no dilution. For high throughput scaling, library preparation reaction volumes and dilutions were miniaturized utilizing acoustic liquid handling (https://protocols.io/view/artic-neb-tagmentation-protocol-high-throughput-wh-bt66nrhe). Briefly, 3 µl of total nucleic acid was used as input for a randomly primed cDNA synthesis reaction. This cDNA served as input for 30 cycles of amplification with ARTIC V3 primers (primer sequences available at https://github.com/artic-network/artic-ncov2019), and was then diluted 1:100 before tagmentation. Adaptor tagmentation was performed using homebrew Tn5, and 8 cycles of index PCR was performed using unique dual barcode Nextera indices. Final libraries were pooled at equal volumes and cleaned at 0.7x (SPRI: Sample) using SPRIselect beads. The library was sequenced on the Illumina Novaseq SP platform in a paired-end 2 x 150 cycle run.

A subset of initial samples were library prepared using the Tailed Amplicon Sequencing V.2 with only primer pairs 71-84 of the ARTIC V3 primers to tile all of the S gene. Final libraries were sequenced by paired-end 2 x 150bp sequencing on an Illumina NovaSeq platform.

### SARS-CoV-2 consensus genome generation

SARS-CoV-2 consensus genomes were generated from raw FASTQ files using the same bioinformatic processes and parameters as defined in (Peng et al. 2021). Base calls were only made at sites with at least 10x high quality read depth, and unambiguous calls were only made if 90% or more of reads at a site specified one particular nucleotide. Viral genomes were uploaded to GISAID (Elbe and Buckland-Merrett 2017), and to NCBI Genbank (Clark et al. 2016) if they had at least 27,500nt (92%) genome coverage with unambiguous base calls and less than 50 non-N ambiguous base calls.

### Phylogenetic dataset collation and analysis

To provide phylogenetic context for viral genomes sampled from Humboldt County, SARS-CoV-2 genome sequences representing global viral diversity were downloaded from GISAID (Shu and McCauley 2017; Elbe and Buckland-Merrett 2017). All phylogenetic analyses to support public health genomic surveillance and to generate figures for this paper were conducted using the Nextstrain tool suite, described in (Hadfield et al. 2018). We used the *Augur* pipeline to align the sequences using nextalign v0.1.6, build a maximum likelihood phylogenetic tree with IQTREE v2.0.3 (Minh et al. 2020), and temporally resolve the tree using TreeTime v0.8.1 (Sagulenko, Puller, and Neher 2018). Final trees, which *Augur* also annotates with nucleotide and amino acid changes across the tree, were exported for visualization in *Auspice*, a web-based application which allows interactive exploration of the phylogenetic trees. To label trees with additional demographic and exposure data describing sequenced cases, we utilized the metadata “drag-and-drop” feature within *Auspice*, which allowed tips in the tree to be colored according to additional data fields specified in a tab-delimited file.

Given the wealth of data available from GISAID, viral genomes collected from regions other than Humboldt County were subsampled according to the following scheme. The 853 samples collected from Humboldt County were considered the focal samples. Given the sheer scale of whole genome sequences available in GISAID, sampling non-Humboldt sequences at random was unlikely to provide proper contextualization of where lineages sampled from Humboldt County circulated prior to migration into the county. Rather, subsampling was designed to maintain spatiotemporal diversity while enriching for contextual genome sequences that were genetically similar to Humboldt County focal samples. To do so, we sampled 50 sequences per month from all other Californian counties that were the least genetically diverged from Humboldt County sequences. Genome sequences from other states in the US were sampled at a rate of 50 genomes per month, again while enriching for sequences that were genetically more similar to sequences sampled from Humboldt County. Finally, to ensure proper rooting and global clade structure of the tree, 5 sequences were sampled per month from each of 6 global regions (Africa, South America, North America (excluding the US), Europe, Asia, and Oceania). The workflow describing the subsampling scheme can be found at https://github.com/alliblk/ncov-humboldt. Genomes used in the analysis had complete date information specifying the year, month and day of sample collection, and had at least 90% genome coverage with non-ambiguous base calls.

### Inference of introduction events to Humboldt County

The geographic migration history between locations was inferred across the tree using the discrete trait analysis method within TreeTime (Sagulenko, Puller, and Neher 2018). Under the phylogeographic model, inferred ancestral viruses (internal nodes within the tree) are annotated with the set of geographic areas where they may possibly have circulated and the probabilities of those states. We defined a migration event into Humboldt County as occurring if a parent node was inferred to circulate with greater than 50% probability anywhere outside of Humboldt County, and the child node was inferred with greater than 50% probability to circulate within Humboldt County. Migration events were considered to seed a discrete introduction into Humboldt County if viruses sampled from individuals residing within Humboldt County were descended from the internal node inferred to circulate within Humboldt County. Because discrete trait phylogeographic models treat sampling intensity as information about the underlying pathogen population size in a deme (Lemey et al. 2009), a phylogeographic analysis with a highly overrepresented deme, such as we have performed here, will bias the reconstruction to favor earlier introductions and longer circulation times. Given this known source of bias, we consciously did not analyze source-sink dynamics or describe transmission directionality between Humboldt County and other geographic areas.

## RESULTS

### Response to SARS-CoV-2 in Humboldt County

Beginning in February 2020, the HCPHL sent COVID-19 testing requests to the Centers for Disease Control and Prevention in Atlanta, Georgia. The first positive case of SARS-CoV-2 in Humboldt County occurred late February 2020. Subsequently, HCPHL started onsite diagnostic testing via quantitative Polymerase Chain Reaction (qPCR) the beginning of March 2020. The first case detected onsite by HCPHL occurred mid-March 2020. The HCPHL capacity and testing strategy evolved over the pandemic, scaling from 10 samples per day at the start of the pandemic to 350 samples per day. By March 2021, the laboratory had screened 35,499 possible cases and detected 1,449 (4.1%) PCR-confirmed SARS-CoV-2 infections (Figure 1). During this time period, two major outbreaks occurred, as described below.

**Figure 1:**
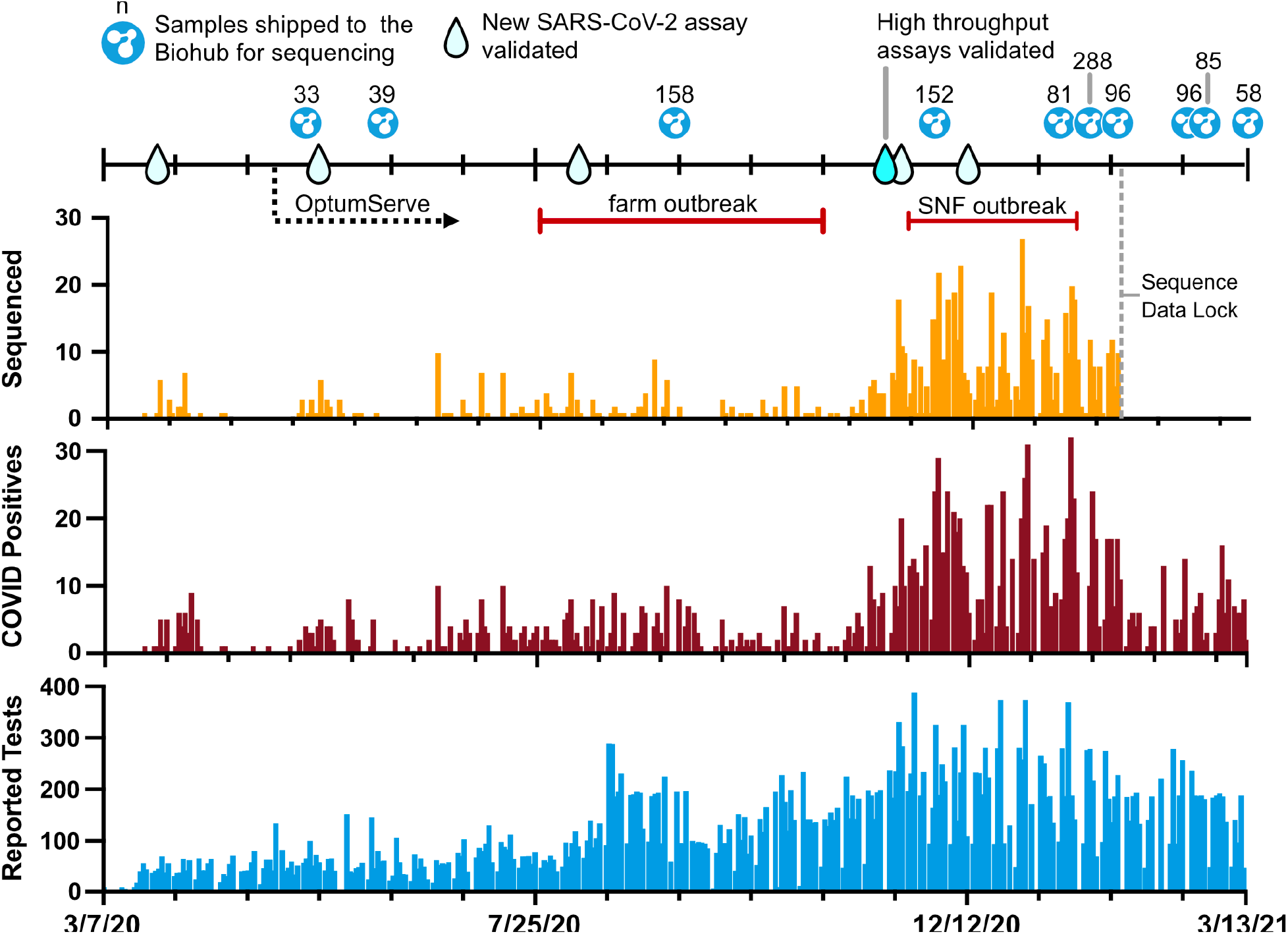
Overview and evolution of testing and sequencing for SARS-CoV-2 in Humboldt County. The blue bar chart indicates the number of tests performed in Humboldt County by the HCPHL by day since the start of testing through the time of writing on March 13, 2021. The maroon bar chart indicates the number of qPCR-positive SARS-CoV-2 cases detected in Humboldt County through time. The orange bar chart indicates the number of viral consensus genome sequences generated from diagnostic specimens over time. Major changes to testing infrastructure are indicated with droplet icons. Shipments for viral genome sequencing are indicated with the CZ Biohub logo.

Like many county health laboratories, HCPHL lacked the resources and infrastructure to conduct whole genome sequencing of SARS-CoV-2 to support surveillance and outbreak response efforts. However, HCPHL established a partnership with the Chan Zuckerberg Biohub’s COVID Tracker Project, a project dedicated to whole genome sequencing and genomic epidemiologic interpretation in support of public health departments throughout California. The HCPHL sought sequencing support initially to assist with case investigations and to monitor for possible viral evolutionary changes that could affect diagnostic assays. The partnership was initiated in May 2020 and enabled Humboldt County to perform genomic surveillance for SARS-CoV-2 throughout the majority of the pandemic, including monitoring for variants of concern later on. A total of 1,086 SARS-CoV-2 positive specimens were sent for sequencing during the time period described (Figure 1).

### Most introductions of SARS-CoV-2 to Humboldt County appear self-limiting or highly-contained

The first sequenced genome from Humboldt County resulted from a sample collected in late March 2020. Between then and the sequence data lock for this manuscript on January 28, 2021, 853 viral whole genome sequences greater than 27,000nt in length were generated from the 1,086 samples sent to CZ Biohub (78.5%). We conducted a phylogeographic analysis of these 853 sequences alongside 1800 contextual SARS-CoV-2 whole genomes, primarily from other counties in California (n=1527), and including sequences from other global regions as well (see Methods for a description of the subsampling scheme).

From our analysis we inferred that there were 100 discrete introductions of SARS-CoV-2 into Humboldt County (Supplemental Figures 1 and 2). The majority of these introduction events led to limited sequenced transmission (Figure 2, Supplemental Figure 1). Of the 100 introduction events, 52 introduction events were singletons, meaning that post-introduction transmission was sufficiently limited that we sequenced only one virus from the introduced lineage. Only 11 introduction events yielded a clade containing more than 10 sequenced infections, with PANGO lineage B.1.311 and B.1.243 strains accounting for the largest number of events (n=222 and 47, respectively (Figure 2)). While not all SARS-CoV-2 positive specimens were sequenced, lineages that transmit for longer periods of time within Humboldt County, or that contribute to large outbreaks, are more likely to yield multiple sequenced isolates. These data suggest that the majority of introductions into Humboldt County were highly contained or self-limiting, while a small number (<10%) of introductions resulted in extended onward transmission within the county.

**Figure 2.**
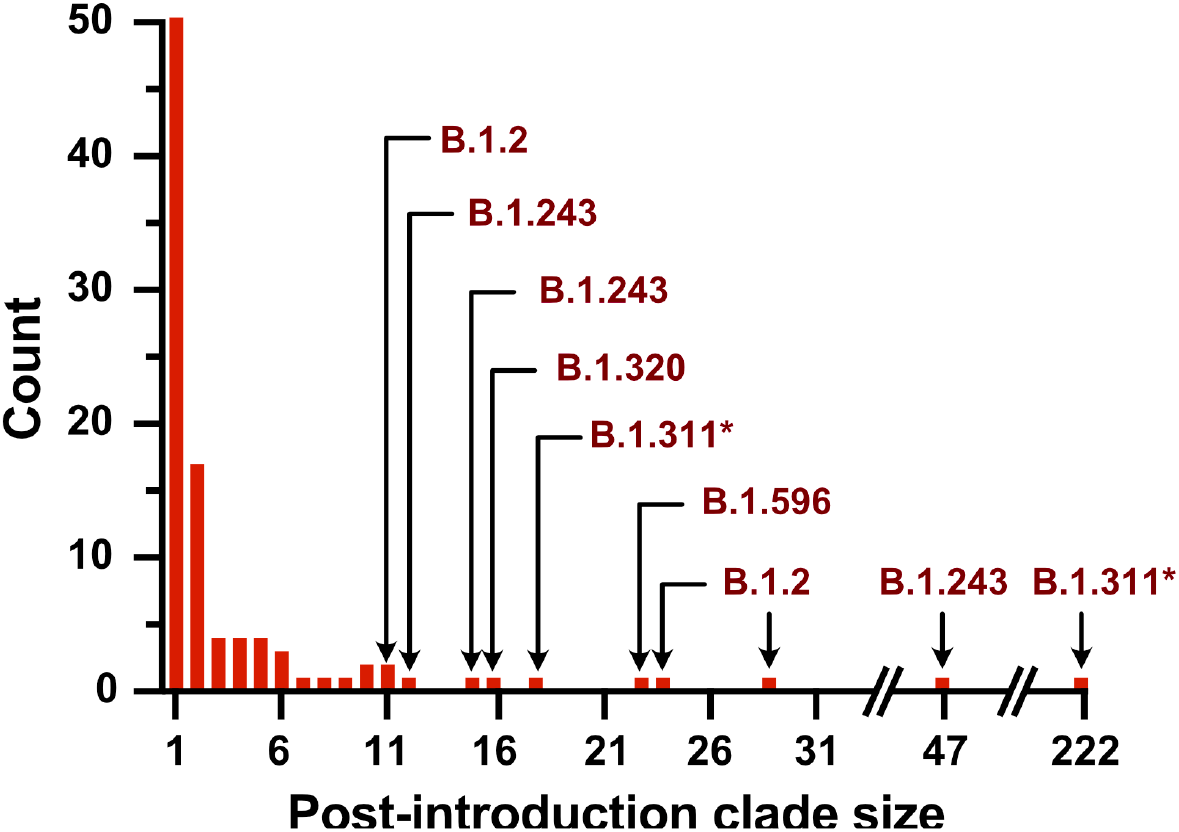
Histogram indicating the number of sequenced viruses grouping within distinct lineages introduced to Humboldt County over the course of the pandemic. For those introductions that resulted in greater than 10 post-introduction events, the PANGO lineage is indicated. While the majority of introductions were limited to themselves, the largest post-introduction clade size was 222 events. *The parental B.1.311 lineage in this case gave rise to additional de novo mutations in Spike in some downstream members of this clade, including N501Y, and T95I.

### Genomic surveillance validates and strengthens contact tracing during a farm-associated outbreak

In late-July 2020, multiple laboratory-confirmed cases of SARS-CoV-2 infections were identified in individuals living in employee housing associated with a commercial farm. The first four cases sought testing at the OptumServe community-testing center in late-July 2020. Case-contact interviews revealed that two of the cases were roommates, and that there was extensive indoor contact with a team lead who was symptomatic. The employee housing at the farm represented a high-contact congregate living facility, and 68 fellow employees were considered close contacts of cases and were ordered to quarantine. An additional 53 individuals who were not employees at the facility were also reported as close contacts and requested to quarantine. DHHS-PH encouraged exposed individuals who developed symptoms to seek testing at either the OptumServe location or at the local hospital, to reduce the burden of travel. Between July 2020 and October 2020, 76 laboratory-confirmed cases were linked to the farm-associated outbreak; 61 cases occurred among employees of the facility and 15 cases occurred among members of the broader community.

Genomic surveillance was initiated by DHHS-PH to characterize the outbreak and guide public health interventions. In total, 37 whole genome sequences were generated that were linked to this outbreak. The outbreak clade was defined by two substitutions: C18744T and G25699T, with additional diversity accruing within the outbreak clade over time (Figure 3). Twenty-nine sequences were generated from diagnostic specimens collected from positive employees at the farm, one sequence came from a non-employee reporting an epidemiologic link to the farm, and seven sequences came from individuals testing positive in the community with no reported link to the farm.

**Figure 3.**
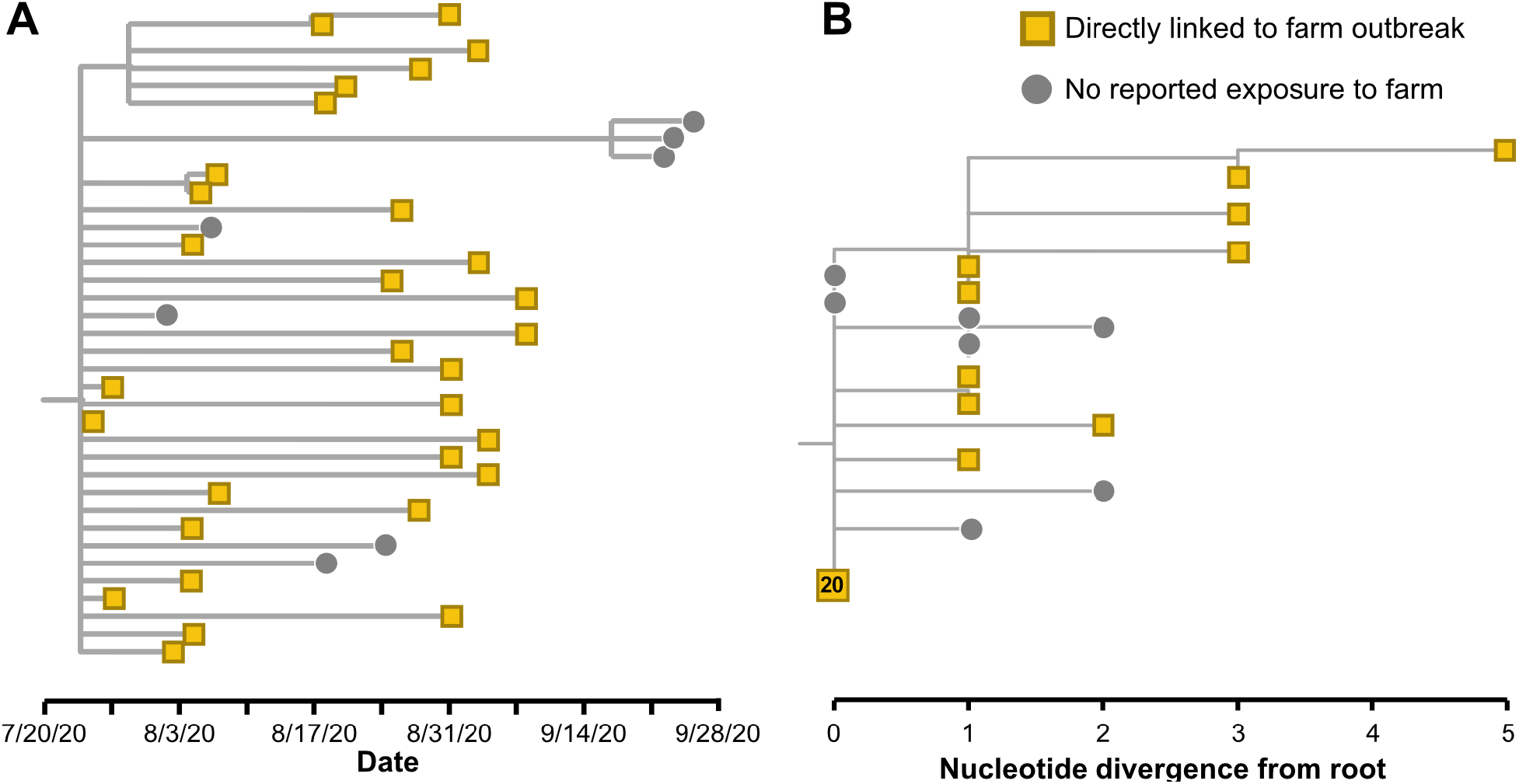
(A) Temporally-resolved phylogenetic tree showing the farm-associated outbreak clade. Viruses collected from individuals reporting an epidemiologic link to the farm are indicated as yellow squares and individuals testing positive within the community with no reported connection with the farm are as grey circles. (B) Maximum likelihood reconstruction of the same farm-associated outbreak clade. Genome sequences that are identical are dispersed along the y-axis at the same internal node. Twenty sequences were identical and are indicated as a single collapsed node on the tree.

Whole genome sequencing helped support contact tracing efforts during this outbreak in various ways. Firstly, pathogen genomic data clearly linked individuals with reported farm exposures to the outbreak (Figure 3), thereby validating information collected in case interviews. Secondly, genomic data indicated where follow-up case investigations were necessary. For example, some individuals did not report farm-associated exposures during their initial case investigation, but had SARS-CoV-2 genome sequences that clustered closely with sequences sampled from the outbreak, linking their infection to the farm. Follow-up investigations for such cases generally revealed an epidemiologic link or exposure to the farm that had not been previously reported, suggesting previously unidentified exposures and transmission chains. Thirdly, some individuals in the community with SARS-CoV-2 cases reported multiple possible exposures, including recent travel and epidemiologic links to the farm outbreak. In these scenarios, viral genomes enabled investigators to determine which exposure had resulted in transmission, and identify other individuals who were at risk from the same exposure event. Finally, genomic data enabled investigators to attribute community cases of SARS-CoV-2 without reported farm exposure to spillover events from the farm outbreak (Figure 3B). Thus, inferences from genomic data allowed epidemiologists to more accurately determine when the outbreak had ended and the extent of forward transmission in the surrounding community.

This outbreak was one of the first large, localized outbreaks that occurred in Humboldt County, and the experience shaped the public health response going forward. This outbreak demonstrated that rapid SARS-CoV-2 transmission could occur within congregate living facilities housing young and active individuals, and that such outbreaks could seed transmission within the wider community. In response, DHHS-PH implemented a specific testing protocol provided by the State to surveil and respond to outbreaks in congregate settings. Under this regime, individuals working or residing in congregate settings were tested for SARS-CoV-2 weekly as surveillance testing. If a positive test was confirmed, this triggered response testing. Contact tracing was initiated and staff and residents were tested twice weekly (if possible) until a two-week period of time with no additional positive tests had elapsed. Additionally, DHHS-PH devoted additional resources to working with congregate living facilities to reduce transmission risk, such as working with commercial agricultural settings to break apart housing into smaller pods, and providing more rapid access to testing.

### Detection of de novo N501Y emergence within a skilled nursing facility outbreak

Like many parts of the United States, Humboldt County experienced a surge in COVID-19 cases during the late-fall and early-winter of 2020. Levels of community transmission were higher, increasing the probability of introduction into congregate living settings at high risk for outbreaks. As part of standard testing procedures for congregate settings, a SARS-CoV-2-positive specimen was collected from a staff member working in a skilled nursing facility, referred to here as Facility A. An outbreak was declared in late-November, 2020, which led to increased testing and contact tracing as per congregate setting protocols described above. Over the course of the outbreak, all 72 residents of the facility tested positive for SARS-CoV-2 (100% attack rate), and 28 of the 98 staff members tested positive (28.6% attack rate). Thirteen residents of the facility died. No mortality was observed amongst infected staff. By mid-January, 2021, no additional positive cases from Facility A had been recorded for two weeks, and the outbreak was declared over.

Routine genomic surveillance of SARS-CoV-2 infections occurring within Humboldt County throughout the fall provided crucial context for understanding the source of this outbreak. In the lead-up to the outbreak in Facility A, a subset of community-associated SARS-CoV-2 cases had been sequenced as part of ongoing genomic surveillance (Figure 4A, grey tips). To identify how SARS-CoV-2 may have been introduced into Facility A, 89 out of the 100 positive specimens collected from staff and residents of Facility A were sequenced. The genome sequence from the postulated index case was more closely related to community cases of SARS-CoV-2 detected in late November 2020 than to sequences collected from other staff and residents at Facility A (Figure 4A). Indeed, their viral genome sequence was diverged from the initial strain that circulated in Facility A by four nucleotide mutations, despite being sampled only one week before the earliest cases reported at Facility A, suggesting that this staff member did not seed the outbreak. The genomic data thus supported an alternative mechanism of introduction into Facility A than was postulated given contact tracing data.

**Figure 4.**
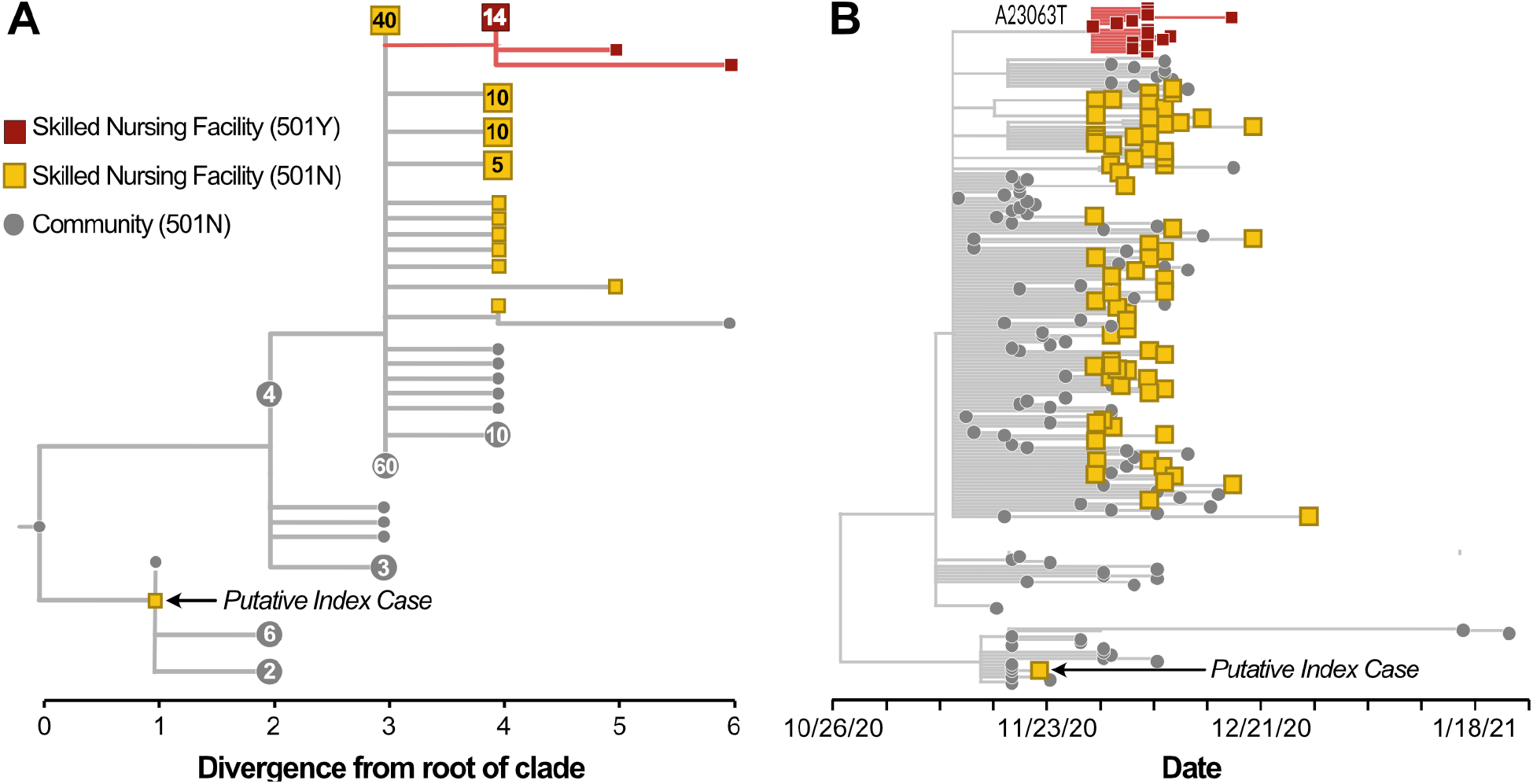
(A) Genetic divergence tree indicating cases within the community (grey tips) and in the skilled nursing facility that had the wildtype N at site 501 in Spike (square yellow tips), and the emergent clade with a Y at site 501 (square maroon tips). The nucleotide substitution yielding the amino acid substitution is annotated on the tree. Square tips represent cases among either the staff or resident population at the SNF, while circular tips represent cases within the community. Large clades of identical genomes are collapsed, with either a square or circular tip, and are annotated with the number of identical genomes that the collapsed tip represents. The staff member that was the putative index case given contact tracing information is annotated. (B) Temporally-resolved phylogenetic tree showing cases sampled from either the SNF (either residents or staff, squares) or the broader community (grey circles). The putative index case is annotated. While the community-associated lineage continued to circulate and was sampled in late-January 2021, the SNF-associated clade was not detected after the end of December 2020.

Genomic surveillance of the outbreak in Facility A also revealed the emergence and eradication of a lineage that had a *de novo* asparagine to tyrosine substitution in site 501 of the Spike protein (N501Y). This particular amino acid substitution is associated with increased binding affinity for the human ACE2 receptor (Starr et al. 2020), which could increase transmissibility. Notably, N501Y is present in three “variant of concern” lineages: B.1.1.7 (Davies et al. 2021), B.1.351 (Tegally et al. 2021), and P.1 (Sabino et al. 2021). Of 89 samples sequenced during this outbreak, 16 samples shared an N501Y substitution (Figure 4, indicated in maroon). The first sample containing this N501Y substitution was collected in late-November, 2020. Within this clade, 14 of the 16 sequences differed from the primary outbreak strain in Facility A by only the A23063T substitution that yielded the N501Y change. Two sequences showed additional diversity (Figure 4A). One virus had an additional C to T substitution at site 19273 which yielded a proline to serine substitution at site 1936 in ORF1b. A second sample showed two C to T mutations at sites 10582 and 23647.

Ongoing community genomic surveillance of positive cases verified eradication of this lineage from Humboldt County. Between the beginning of January 2021 and the sequence data lock for this manuscript, 233 sequences collected from SARS-CoV-2 cases in Humboldt County were generated, yet none of these samples grouped with, or were descended from, the 501N or 501Y lineages that circulated in Facility A. Two cases sampled in January 2021 had viral genome sequences that were descended from the lineage that circulated in the community (Figure 4B). These data suggest that intervention efforts effectively eradicated Facility A’s outbreak lineages even as transmission within the wider community occurred.

## DISCUSSION

Here we have described the development of increased laboratory capacity and use of genomic surveillance to support the public health response to COVID-19 in a local health department. While genomic surveillance data have frequently been used to understand geographical patterns of infectious disease transmission, our work here demonstrates the utility of pathogen genomic sequence data for supporting actionable public health activities, such as contact tracing and outbreak response, within a rural county locale. Due to the rural nature of Humboldt County, there are generally longer turnaround times between sample collection and a test result because of the need to transport samples longer distances. Sample transport usually adds several days to the time necessary to receive a test result, and these delays can affect the ability to respond locally in a timely manner. To mitigate the impact of these issues, the HCPHL focused on modernizing their public health surveillance capabilities, including scaling up testing infrastructure and integrating viral genomic surveillance into response efforts.

These laboratory investments likely contributed significantly to DHHS-PH’s epidemiologic response efforts, thereby limiting community spread for many months. Combining traditional contact tracing with phylogenetic analysis of viral sequence data facilitated rapid identification and response in three important ways. First, sustained community transmission within the county could be actively monitored, instead of relying solely on state-wide or national data. Had DHHS-PH been reliant on state-wide or national data, several key opportunities to control local transmission may have been missed. Second, the ability to confirm findings from case-contact interviews with genomic data allowed determination of which exposure event had resulted in transmission given multiple possible exposures, providing direct feedback on the efficacy of the overall response efforts. Finally, these new capabilities allowed early identification of introductions or emergence of variants of concern for immediate monitoring, containment, and local elimination.

The genomic surveillance findings in Humboldt County suggest that the majority of introductions into the county resulted in limited onward transmission and successful local control with only 11 (11%) of documented introduction events resulting in greater than 10 sequenced cases. Notably, these results are likely conservative due to the directionality of the bias in the phylogeographic reconstruction due to the large number of samples incorporated from Humboldt County. Additional sequence data from other geographic areas would likely interdigitate with, and potentially break apart, some of the clades inferred to circulate within Humboldt County. In this case, the number of discrete introductions into Humboldt County would be greater, and the amount of onward transmission observed post-introduction would decrease. Thus our estimate of 100 distinct introductions of SARS-CoV-2 to Humboldt County should be viewed as the minimal estimate of introduction frequency, and lineages may be even more self-limiting than observed in this study. Our analyses also revealed how epidemiologic dynamics shifted from iterative introductions resulting in limited community spread to large congregate setting outbreaks as the overall number of cases increased in the summer and fall of 2020. Congregate setting outbreaks were observed that were contained to the facility (SNF) as well as seeding broader viral spread within the community (farm).

Using this genomic surveillance system we detected the emergence of a viral strain with a *de novo* N501Y substitution in the Spike protein. While the phenotypic characteristics of this substitution within this specific genetic context are not known, DHHS-PH’s response to this emergence event appears to have resulted in eradication of this lineage. This was feasible because a large proportion of positive diagnostic specimens collected from the surrounding community was consistently being sequenced. Monitoring these community samples in addition to specimens collected from the SNF where the substitution arose demonstrated that the lineage had been successfully contained to the facility and did not spill over and transmit within the broader community. To our knowledge, this is the first instance of a local health department within the United States detecting an emergent mutation of interest and documenting its eradication with genomic surveillance.

Genomic surveillance is still considered an advanced technique in public health and is often more accessible at higher levels of jurisdictional authority or in higher resource urban settings. This thinking has less to do with implementation of laboratory protocols for conducting sequencing, which are increasingly accessible to most public health microbiology labs with molecular capabilities. Rather, this issue arises due to the unique challenges associated with genomic data management, analysis, and interpretation (Armstrong et al. 2019; Black et al. 2020). However, our experience in Humboldt County shows the value in supporting genomic surveillance capacity at the local level and integrating it with event- and indicator-based surveillance efforts. Public health agencies at the local level are typically the frontline response to local public health issues. Thus, improving these agencies’ ability to generate, analyze and interpret genomic data rapidly makes the data more actionable for public health decision making. To support this objective the California COVID Tracker Project is transitioning from providing genomic epidemiology as a service to supporting development of in-house genomic surveillance capacity for local public health departments. This effort includes hands-on training, education, and lowering analysis barriers by developing open-source software tools. Such partnerships can provide invaluable technology and knowledge transfer to local public health departments, which will foster timely, representative, flexible, and responsive deployment of genomic epidemiology for surveillance and outbreak response based on local needs and priorities. If facilitated by a sustained increase in government funding, these partnership and capacity building models have the potential to facilitate genomic surveillance programs at the county level and modernize public health surveillance across all levels of jurisdictional authority.

## Data Availability

The Augur and Auspice components of the Nextstrain workflow are publicly available at https://github.com/nextstrain/. The SARS-CoV-2-specific Nextstrain phylogenetic pipeline is publicly available at https://github.com/nextstrain/ncov. The profile that specifies our particular parameters for this workflow is available at https://github.com/alliblk/ncov-humboldt. Scripts used in data analysis are also available at https://github.com/alliblk/ncov-humboldt. Whole genome sequences generated by CZ Biohub are available within GISAID and NCBI GenBank.

## DATA AND CODE AVAILABILITY

The *Augur* and *Auspice* components of the Nextstrain workflow are publicly available at https://github.com/nextstrain/. The SARS-CoV-2-specific Nextstrain phylogenetic pipeline is publicly available at https://github.com/nextstrain/ncov. The profile that specifies our particular parameters for this workflow is available at https://github.com/alliblk/ncov-humboldt. Scripts used in data analysis are also available at https://github.com/alliblk/ncov-humboldt. Whole genome sequences generated by CZ Biohub are available within GISAID and NCBI GenBank; accession numbers are given in Supplemental Table 1.

## ACKNOWLEDGEMENTS

The authors would like to recognize the tireless efforts of Humboldt County’s COVID-19 Investigation Task Force Leads, Angela Winogradov and Erica Dykehouse. The success of the county’s COVID-19 response is due in no small part to the dedication and passion evident in their work. We also thank the EOC Health & Wellness Branch Director Megan Blanchard, COVID Investigations Group Supervisor Helen Culver, and Public Health Communicable Disease Supervisor Hava Phillips for their exemplary leadership and dedication to their community as demonstrated over the course of the COVID-19 pandemic. We would also like to thank our Public Health Director, Michele Stephens, our Health Officers Dr. Terry Frankovich, Dr. Josh Ennis and Dr. Ian Hoffman, for their sacrifice, support and leadership for the duration of the response. We wish to recognize the valued members of our community who have volunteered their time to assist the DHHS-PH COVID-19 Response with case investigations, contact tracing, and data management. We also want to thank the amazing HCPHL staff for their dedication to Public Health, their scientific expertise and endless hours spent testing COVID-19 samples during this pandemic. Furthermore, we recognize our DHHS-PH COVID response team that have dedicated countless hours of planning, response and support for our community. The authors kindly thank Dr. Cristina M Tato for her support and feedback on this manuscript from conceptualization of the paper through to writing and editing the manuscript. The Covid Tracker Project is a team effort, and all members provide critical contributions to a system that has facilitated genomic surveillance across California throughout the duration of the outbreak. In particular, we would like to thank our clinical research coordinator Maira Phelps, who has worked tirelessly to coordinate specimen shipments to CZ Biohub from all of our county partners; Shannon Axelrod and Phoenix Logan, who developed the LIMS system for COVID Tracker, Dr. David Dynerman, who helped build the frontend visualization portal for COVID Tracker, Tony Tung, who developed and maintained an enormous automated system for running Nextstrain builds, Dr. Sidney Bell, who provided essential expertise and technical knowledge in the inception, development, and continued growth of this project, and Dr. Tiffany J. Chen for her leadership and work to integrate this genomic surveillance project into the broader scientific ecosystem. Finally, we are incredibly grateful to the numerous labs that have been sequencing and depositing genomic sequences and metadata to public repositories throughout the pandemic, thereby enabling other groups to contextualize their own data and understand transmission within their jurisdictions.

---

**Supplemental Figure 1:**
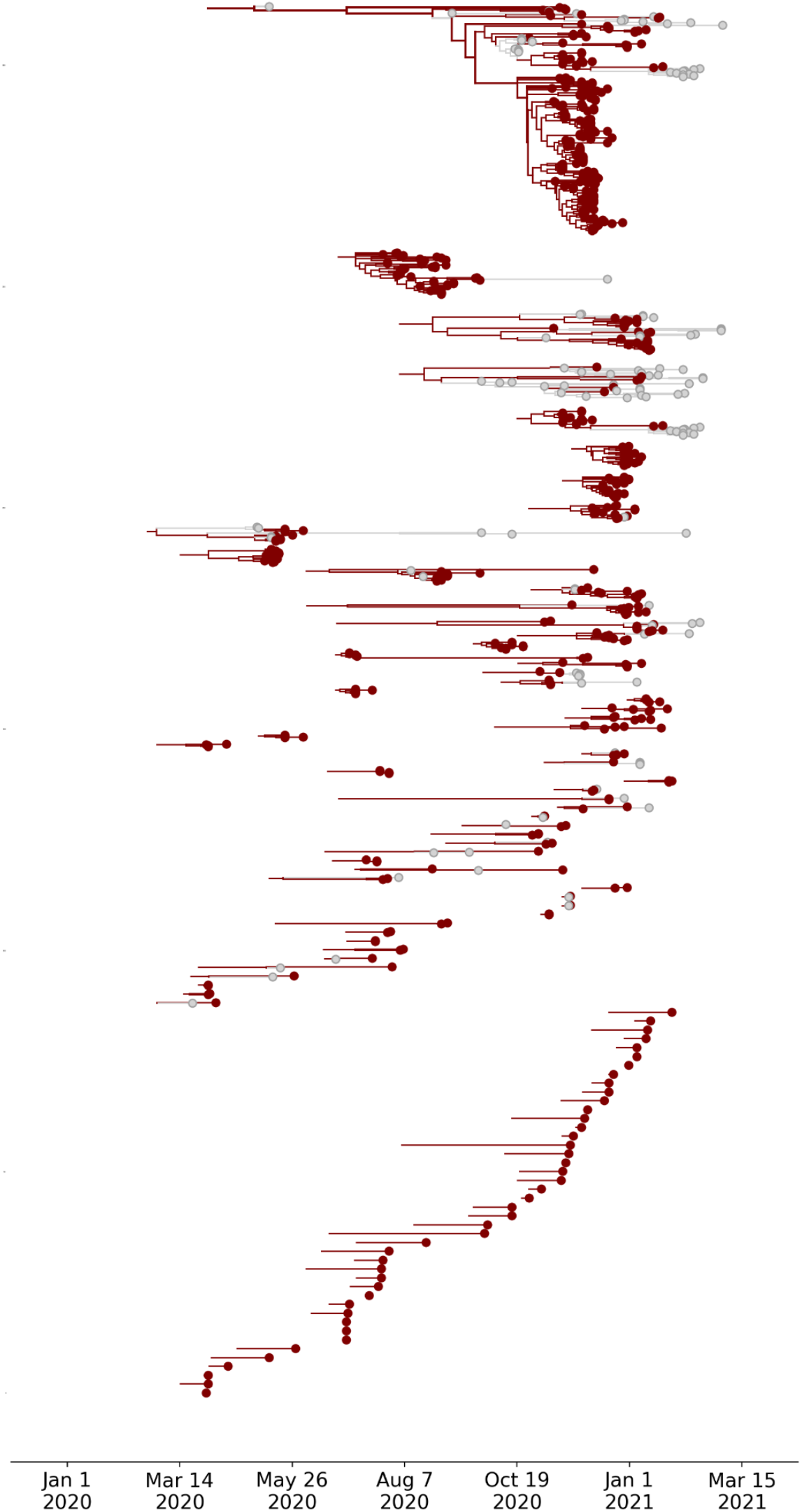
Phylogenetic clades for 100 discrete lineages of SARS-CoV-2 introduced to Humboldt County. Each clade represents a unique introduction of the virus to the county, and the length of maroon branches indicate transmission duration within Humboldt County. Grey tips and branches represent descendants of that lineage that circulated outside of Humboldt County. Most introductions lead to limited transmission within Humboldt County, as evidenced by minimal genetic diversity of a clade sampled within the county.

**Supplemental Figure 2:**
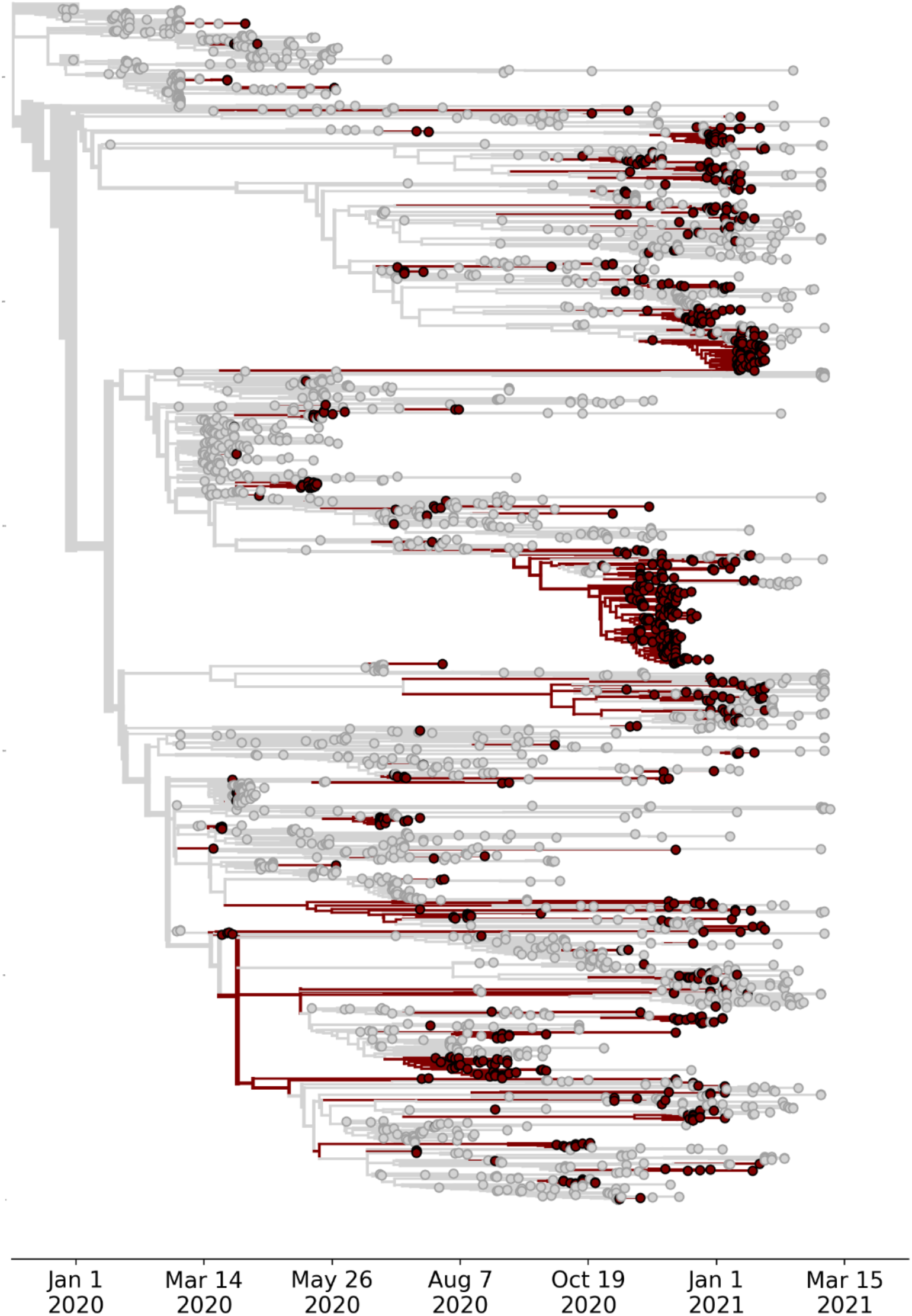
Temporally-resolved phylogenetic tree of 2653 SARS-CoV-2 genomes, 853 of which were sampled from Humboldt County. Tips sampled from Humboldt County are indicated in maroon and tips sampled from other locations are colored in grey. Branches colored in maroon indicate inferred circulation within Humboldt County, and branches in grey indicate inferred transmission outside of Humboldt County.

