## Supplementary Information for "Using genomic epidemiology of SARS-CoV-2 to support contact tracing and public health surveillance in rural Humboldt County, California"

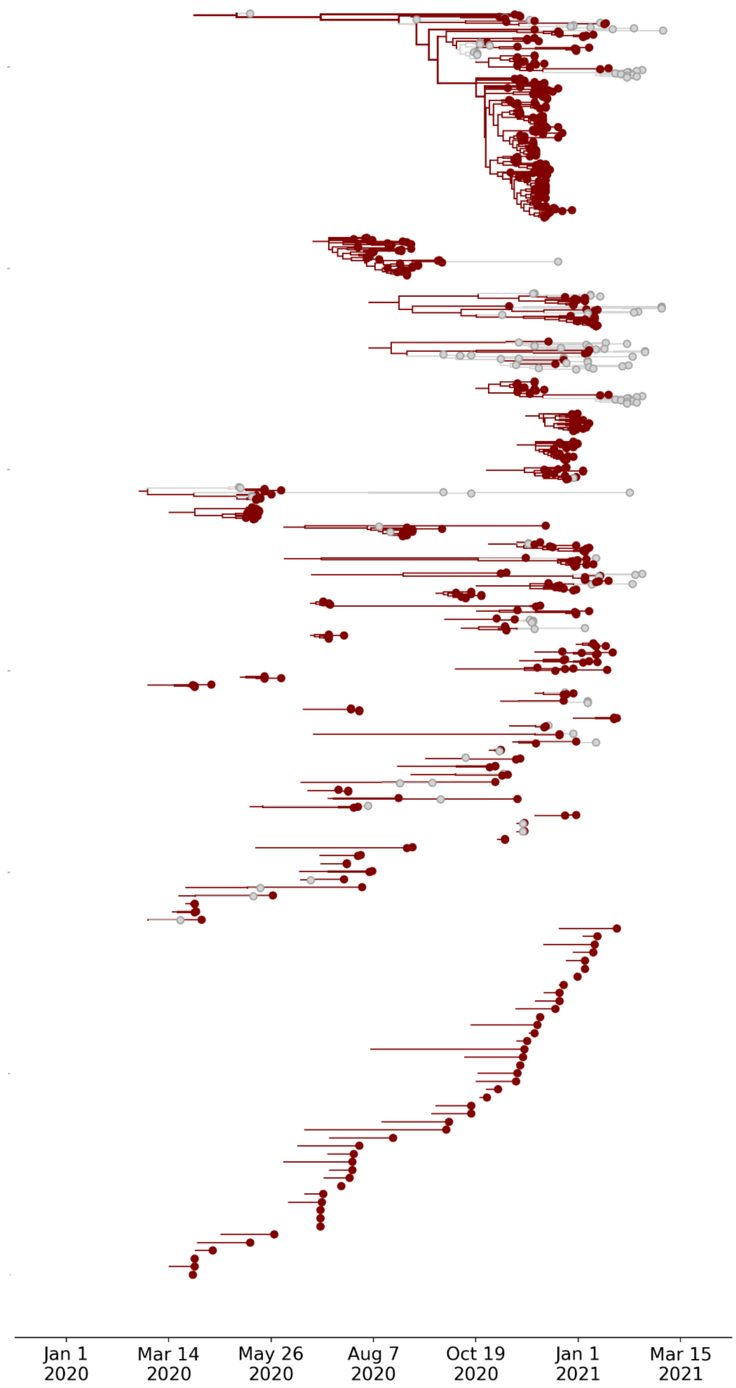


Supplemental Figure 1: Phylogenetic clades for 100 discrete lineages of SARS-CoV-2 introduced to Humboldt County. Each clade represents a unique introduction of the virus to the county, and the length of maroon branches indicate transmission duration within Humboldt County. Grey tips and branches represent descendents of that lineage that circulated outside of Humboldt County. Most introductions lead to limited transmission within Humboldt County, as evidenced by minimal genetic diversity of a clade sampled within the county.

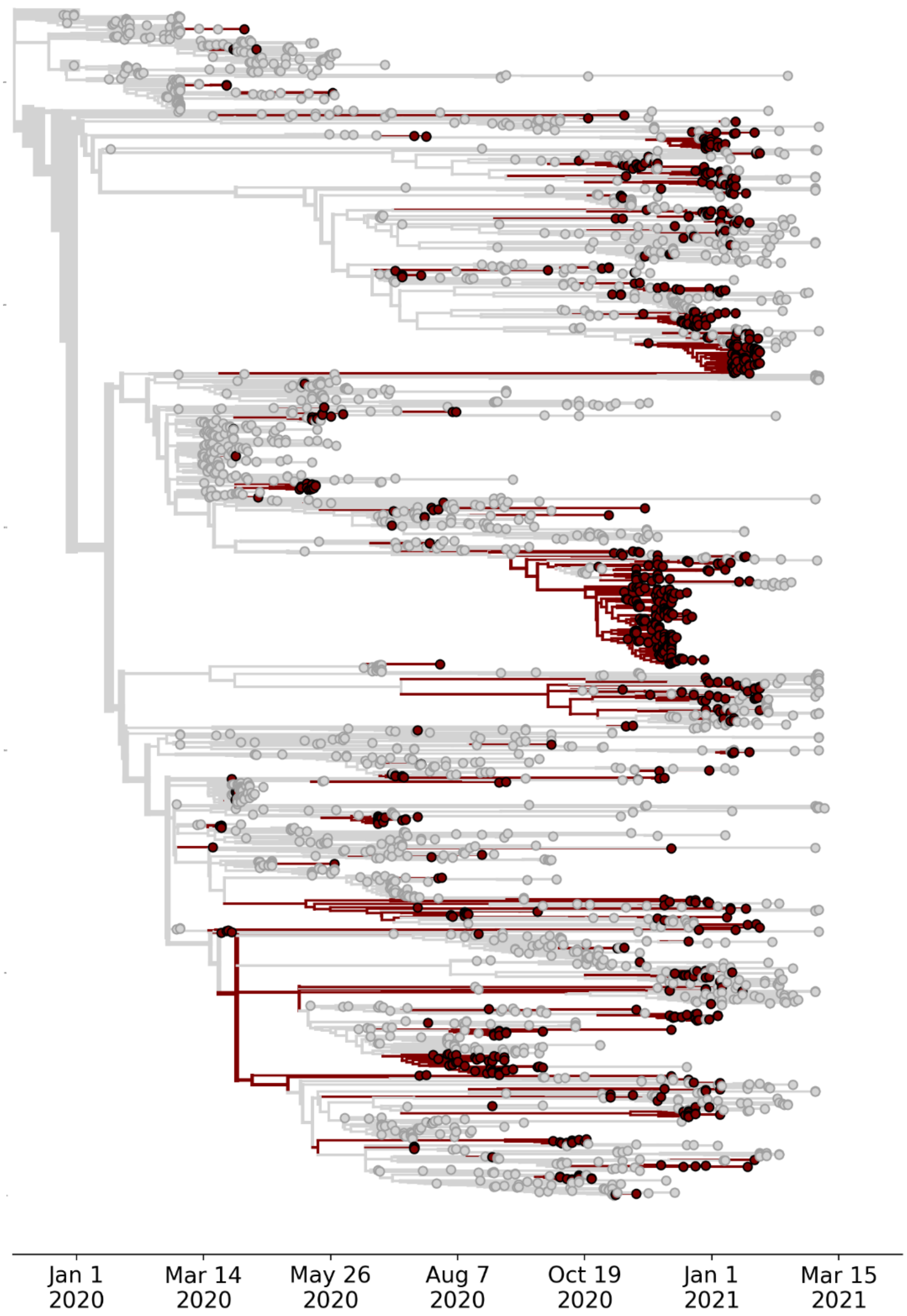


Supplemental Figure 2: Temporally-resolved phylogenetic tree of 2653 SARS-CoV-2 genomes, 853 of which were sampled from Humboldt County. Tips sampled from Humboldt County are indicated in maroon and tips sampled from other locations are colored in grey. Branches colored in maroon indicate inferred circulation within Humboldt County, and branches in grey indicate inferred transmission outside of Humboldt County.

Supplemental Table 1: Sequence data accession numbers. All sequences were submitted to GISAID as they were generated, and accession numbers are given. At the end of the study sequences were also deposited into NCBI GenBank. Some sequences accepted by GISAID were rejected by NCBI. In these cases, the GenBank accession number is given as “NA”.

| **Sample_ID** | **GISAID_accession** | **GenBank_accession** |
|---|---|---|
| USA/CA-CZB-10785/2020 | EPI_ISL_583046 | MW276323 |
| USA/CA-CZB-10786/2020 | EPI_ISL_583047 | MW276324 |
| USA/CA-CZB-10788/2020 | EPI_ISL_583048 | MW276325 |
| USA/CA-CZB-10789/2020 | EPI_ISL_583049 | MW276326 |
| USA/CA-CZB-10790/2020 | EPI_ISL_583050 | MW276327 |
| USA/CA-CZB-10791/2020 | EPI_ISL_583051 | MW276328 |
| USA/CA-CZB-10792/2020 | EPI_ISL_583052 | MW276329 |
| USA/CA-CZB-10793/2020 | EPI_ISL_583053 | MW276330 |
| USA/CA-CZB-10794/2020 | EPI_ISL_583054 | MW276331 |
| USA/CA-CZB-10795/2020 | EPI_ISL_583055 | MW276332 |
| USA/CA-CZB-10796/2020 | EPI_ISL_583056 | MW276333 |
| USA/CA-CZB-10797/2020 | EPI_ISL_583057 | MW276334 |
| USA/CA-CZB-10798/2020 | EPI_ISL_583058 | MW276335 |
| USA/CA-CZB-10799/2020 | EPI_ISL_2659414 | MZ842558 |
| USA/CA-CZB-10800/2020 | EPI_ISL_2659492 | MZ842557 |
| USA/CA-CZB-10801/2020 | EPI_ISL_583059 | MW276336 |
| USA/CA-CZB-10802/2020 | EPI_ISL_583060 | MW276337 |
| USA/CA-CZB-10803/2020 | EPI_ISL_2659500 | MZ842556 |
| USA/CA-CZB-10804/2020 | EPI_ISL_583061 | MW276338 |
| USA/CA-CZB-10805/2020 | EPI_ISL_2659209 | MZ842555 |
| USA/CA-CZB-10808/2020 | EPI_ISL_2659371 | MZ842554 |
| USA/CA-CZB-10809/2020 | EPI_ISL_583062 | MW276339 |
| USA/CA-CZB-10810/2020 | EPI_ISL_583063 | MW276340 |
| USA/CA-CZB-10811/2020 | EPI_ISL_583064 | MW276341 |
| USA/CA-CZB-10812/2020 | EPI_ISL_583065 | MW276342 |

| | | |
|---|---|---|
| USA/CA-CZB-10813/2020 | EPI_ISL_583066 | MW276343 |
| USA/CA-CZB-10815/2020 | EPI_ISL_583067 | MW276344 |
| USA/CA-CZB-10817/2020 | EPI_ISL_2659405 | MZ842553 |
| USA/CA-CZB-10818/2020 | EPI_ISL_583068 | MW276345 |
| USA/CA-CZB-10819/2020 | EPI_ISL_583069 | MW276346 |
| USA/CA-CZB-10820/2020 | EPI_ISL_583070 | MW276347 |
| USA/CA-CZB-10821/2020 | EPI_ISL_583071 | MW276348 |
| USA/CA-CZB-10822/2020 | EPI_ISL_583072 | MW276349 |
| USA/CA-CZB-10823/2020 | EPI_ISL_583073 | MW276350 |
| USA/CA-CZB-10824/2020 | EPI_ISL_583074 | MW276351 |
| USA/CA-CZB-10825/2020 | EPI_ISL_583075 | MW276352 |
| USA/CA-CZB-10826/2020 | EPI_ISL_583076 | MW276353 |
| USA/CA-CZB-10827/2020 | EPI_ISL_583077 | MW276354 |
| USA/CA-CZB-10828/2020 | EPI_ISL_583078 | MW276355 |
| USA/CA-CZB-10829/2020 | EPI_ISL_2758407 | MZ842552 |
| USA/CA-CZB-10830/2020 | EPI_ISL_583079 | MW276356 |
| USA/CA-CZB-10831/2020 | EPI_ISL_2659486 | MZ842551 |
| USA/CA-CZB-10832/2020 | EPI_ISL_583080 | MW276357 |
| USA/CA-CZB-10833/2020 | EPI_ISL_2659487 | MZ842550 |
| USA/CA-CZB-10834/2020 | EPI_ISL_583081 | MW276358 |
| USA/CA-CZB-10835/2020 | EPI_ISL_583082 | MW276359 |
| USA/CA-CZB-10836/2020 | EPI_ISL_583083 | MW276360 |
| USA/CA-CZB-10837/2020 | EPI_ISL_583084 | MW276361 |
| USA/CA-CZB-10838/2020 | EPI_ISL_2758408 | MZ842549 |
| USA/CA-CZB-10839/2020 | EPI_ISL_583085 | MW276362 |
| USA/CA-CZB-10841/2020 | EPI_ISL_583086 | MW276363 |
| USA/CA-CZB-10842/2020 | EPI_ISL_583087 | MW276364 |
| USA/CA-CZB-10843/2020 | EPI_ISL_583088 | MW276365 |
| USA/CA-CZB-10844/2020 | EPI_ISL_583089 | MW276366 |

| | | |
|---|---|---|
| USA/CA-CZB-10845/2020 | EPI_ISL_583090 | MZ842548 |
| USA/CA-CZB-10846/2020 | EPI_ISL_583091 | MW276367 |
| USA/CA-CZB-10847/2020 | EPI_ISL_583092 | MW276368 |
| USA/CA-CZB-10848/2020 | EPI_ISL_583093 | MW276369 |
| USA/CA-CZB-10849/2020 | EPI_ISL_583094 | MW276370 |
| USA/CA-CZB-10850/2020 | EPI_ISL_583095 | MW276371 |
| USA/CA-CZB-10851/2020 | EPI_ISL_583096 | MW276372 |
| USA/CA-CZB-10852/2020 | EPI_ISL_583097 | MW276373 |
| USA/CA-CZB-10854/2020 | EPI_ISL_583098 | MW276374 |
| USA/CA-CZB-10855/2020 | EPI_ISL_583099 | MW276375 |
| USA/CA-CZB-10856/2020 | EPI_ISL_583100 | MW276376 |
| USA/CA-CZB-10858/2020 | EPI_ISL_583101 | MZ842547 |
| USA/CA-CZB-10860/2020 | EPI_ISL_583102 | MW276377 |
| USA/CA-CZB-10861/2020 | EPI_ISL_583103 | MW276378 |
| USA/CA-CZB-10862/2020 | EPI_ISL_583104 | MW276379 |
| USA/CA-CZB-10863/2020 | EPI_ISL_583105 | MW276380 |
| USA/CA-CZB-10864/2020 | EPI_ISL_2758409 | NA |
| USA/CA-CZB-10865/2020 | EPI_ISL_583106 | MW276381 |
| USA/CA-CZB-10866/2020 | EPI_ISL_583107 | MW276382 |
| USA/CA-CZB-10867/2020 | EPI_ISL_583108 | MW276383 |
| USA/CA-CZB-10868/2020 | EPI_ISL_583109 | MW276384 |
| USA/CA-CZB-10870/2020 | EPI_ISL_583110 | MW276385 |
| USA/CA-CZB-10871/2020 | EPI_ISL_583111 | MW276386 |
| USA/CA-CZB-10872/2020 | EPI_ISL_583112 | MW276387 |
| USA/CA-CZB-10873/2020 | EPI_ISL_583113 | MW276388 |
| USA/CA-CZB-10874/2020 | EPI_ISL_583114 | MW276389 |
| USA/CA-CZB-10876/2020 | EPI_ISL_583115 | MW276390 |
| USA/CA-CZB-10878/2020 | EPI_ISL_583116 | MW276391 |
| USA/CA-CZB-10879/2020 | EPI_ISL_583117 | MW276392 |

| | | |
|---|---|---|
| USA/CA-CZB-10880/2020 | EPI_ISL_583118 | MW276393 |
| USA/CA-CZB-10881/2020 | EPI_ISL_583119 | MW276394 |
| USA/CA-CZB-10885/2020 | EPI_ISL_583120 | MW276395 |
| USA/CA-CZB-10887/2020 | EPI_ISL_583121 | MW276396 |
| USA/CA-CZB-10889/2020 | EPI_ISL_583122 | MW276397 |
| USA/CA-CZB-10891/2020 | EPI_ISL_583123 | MW276398 |
| USA/CA-CZB-10892/2020 | EPI_ISL_583124 | MW276399 |
| USA/CA-CZB-10893/2020 | EPI_ISL_2658689 | NA |
| USA/CA-CZB-10895/2020 | EPI_ISL_583125 | MW276400 |
| USA/CA-CZB-10896/2020 | EPI_ISL_583126 | MW276401 |
| USA/CA-CZB-10897/2020 | EPI_ISL_583127 | MW276402 |
| USA/CA-CZB-10898/2020 | EPI_ISL_583128 | MW276403 |
| USA/CA-CZB-10899/2020 | EPI_ISL_583129 | MW276404 |
| USA/CA-CZB-10901/2020 | EPI_ISL_583130 | MW276405 |
| USA/CA-CZB-10902/2020 | EPI_ISL_583131 | MW276406 |
| USA/CA-CZB-10903/2020 | EPI_ISL_583132 | MW276407 |
| USA/CA-CZB-10904/2020 | EPI_ISL_583133 | MW276408 |
| USA/CA-CZB-10905/2020 | EPI_ISL_583134 | MW276409 |
| USA/CA-CZB-10906/2020 | EPI_ISL_583135 | MW276410 |
| USA/CA-CZB-10907/2020 | EPI_ISL_583136 | MW276411 |
| USA/CA-CZB-10908/2020 | EPI_ISL_583137 | MW276412 |
| USA/CA-CZB-10909/2020 | EPI_ISL_583138 | MW276413 |
| USA/CA-CZB-10910/2020 | EPI_ISL_583139 | MW276414 |
| USA/CA-CZB-10911/2020 | EPI_ISL_583140 | MW276415 |
| USA/CA-CZB-10912/2020 | EPI_ISL_583141 | MW276416 |
| USA/CA-CZB-10913/2020 | EPI_ISL_583142 | MW276417 |
| USA/CA-CZB-10914/2020 | EPI_ISL_583143 | MW276418 |
| USA/CA-CZB-10915/2020 | EPI_ISL_583144 | MW276419 |
| USA/CA-CZB-10916/2020 | EPI_ISL_583145 | MW276420 |

| | | |
|---|---|---|
| USA/CA-CZB-10917/2020 | EPI_ISL_583146 | MW276421 |
| USA/CA-CZB-10918/2020 | EPI_ISL_583147 | MW276422 |
| USA/CA-CZB-10919/2020 | EPI_ISL_583148 | MW276423 |
| USA/CA-CZB-10920/2020 | EPI_ISL_583149 | MW276424 |
| USA/CA-CZB-10921/2020 | EPI_ISL_583150 | MW276425 |
| USA/CA-CZB-10922/2020 | EPI_ISL_583151 | MW276426 |
| USA/CA-CZB-10923/2020 | EPI_ISL_583152 | MW276427 |
| USA/CA-CZB-10924/2020 | EPI_ISL_583153 | MW276428 |
| USA/CA-CZB-10925/2020 | EPI_ISL_583154 | MW276429 |
| USA/CA-CZB-10926/2020 | EPI_ISL_583155 | MW276430 |
| USA/CA-CZB-10927/2020 | EPI_ISL_583156 | MW276431 |
| USA/CA-CZB-10928/2020 | EPI_ISL_583157 | MW276432 |
| USA/CA-CZB-10929/2020 | EPI_ISL_583158 | MW276433 |
| USA/CA-CZB-10930/2020 | EPI_ISL_583159 | MW276434 |
| USA/CA-CZB-10931/2020 | EPI_ISL_583160 | MW276435 |
| USA/CA-CZB-10932/2020 | EPI_ISL_583161 | MW276436 |
| USA/CA-CZB-10933/2020 | EPI_ISL_583162 | MW276437 |
| USA/CA-CZB-10934/2020 | EPI_ISL_583163 | MW276438 |
| USA/CA-CZB-10935/2020 | EPI_ISL_583164 | MW276439 |
| USA/CA-CZB-10936/2020 | EPI_ISL_583165 | MW276440 |
| USA/CA-CZB-10937/2020 | EPI_ISL_583166 | MW276441 |
| USA/CA-CZB-10938/2020 | EPI_ISL_583167 | MW276442 |
| USA/CA-CZB-10939/2020 | EPI_ISL_583168 | MW276443 |
| USA/CA-CZB-10940/2020 | EPI_ISL_583169 | MW276444 |
| USA/CA-CZB-10941/2020 | EPI_ISL_583170 | MW276445 |
| USA/CA-CZB-1278/2020 | EPI_ISL_454636 | MT533233 |
| USA/CA-CZB-1279/2020 | EPI_ISL_454637 | MT533234 |
| USA/CA-CZB-1280/2020 | EPI_ISL_454638 | MT533235 |
| USA/CA-CZB-1281/2020 | EPI_ISL_454639 | MT533236 |

| | | |
|---|---|---|
| USA/CA-CZB-1282/2020 | EPI_ISL_454640 | MT533237 |
| USA/CA-CZB-1283/2020 | EPI_ISL_454641 | MT533238 |
| USA/CA-CZB-14715/2020 | EPI_ISL_738785 | NA |
| USA/CA-CZB-14716/2020 | EPI_ISL_738778 | MW565247 |
| USA/CA-CZB-14717/2020 | EPI_ISL_739011 | MW565246 |
| USA/CA-CZB-14718/2020 | EPI_ISL_2658704 | NA |
| USA/CA-CZB-14721/2020 | EPI_ISL_739489 | MW565245 |
| USA/CA-CZB-14722/2020 | EPI_ISL_739264 | NA |
| USA/CA-CZB-14724/2020 | EPI_ISL_739086 | MW565244 |
| USA/CA-CZB-14725/2020 | EPI_ISL_2659408 | MZ842401 |
| USA/CA-CZB-14727/2020 | EPI_ISL_739297 | MW565243 |
| USA/CA-CZB-14728/2020 | EPI_ISL_2659346 | MZ842400 |
| USA/CA-CZB-14729/2020 | EPI_ISL_739571 | MW565242 |
| USA/CA-CZB-14730/2020 | EPI_ISL_739366 | MW565241 |
| USA/CA-CZB-14731/2020 | EPI_ISL_2659403 | MZ842399 |
| USA/CA-CZB-14733/2020 | EPI_ISL_2659591 | MZ842398 |
| USA/CA-CZB-14734/2020 | EPI_ISL_738988 | MW565240 |
| USA/CA-CZB-14735/2020 | EPI_ISL_2659461 | MZ842397 |
| USA/CA-CZB-14736/2020 | EPI_ISL_739311 | MW565239 |
| USA/CA-CZB-14737/2020 | EPI_ISL_739533 | MW565238 |
| USA/CA-CZB-14739/2020 | EPI_ISL_2659260 | MZ842396 |
| USA/CA-CZB-14740/2020 | EPI_ISL_2659385 | MZ842395 |
| USA/CA-CZB-14741/2020 | EPI_ISL_739177 | MW565237 |
| USA/CA-CZB-14742/2020 | EPI_ISL_738961 | MW565236 |
| USA/CA-CZB-14743/2020 | EPI_ISL_739558 | MW565235 |
| USA/CA-CZB-14744/2020 | EPI_ISL_2659205 | MZ842394 |
| USA/CA-CZB-14745/2020 | EPI_ISL_738530 | MW565234 |
| USA/CA-CZB-14746/2020 | EPI_ISL_2659549 | MZ842393 |
| USA/CA-CZB-14747/2020 | EPI_ISL_2659638 | MZ842392 |

| | | |
|---|---|---|
| USA/CA-CZB-14748/2020 | EPI_ISL_738879 | MW565233 |
| USA/CA-CZB-14749/2020 | EPI_ISL_738756 | MW565232 |
| USA/CA-CZB-14750/2020 | EPI_ISL_739225 | MW565231 |
| USA/CA-CZB-14751/2020 | EPI_ISL_738951 | MW565230 |
| USA/CA-CZB-14752/2020 | EPI_ISL_739148 | MW565229 |
| USA/CA-CZB-14754/2020 | EPI_ISL_738947 | MW565227 |
| USA/CA-CZB-14755/2020 | EPI_ISL_739433 | MW565226 |
| USA/CA-CZB-14756/2020 | EPI_ISL_739530 | MW565225 |
| USA/CA-CZB-14757/2020 | EPI_ISL_739442 | MZ842391 |
| USA/CA-CZB-14758/2020 | EPI_ISL_738953 | MZ842390 |
| USA/CA-CZB-14759/2020 | EPI_ISL_738832 | MW565224 |
| USA/CA-CZB-14760/2020 | EPI_ISL_739052 | MW565223 |
| USA/CA-CZB-14767/2020 | EPI_ISL_739051 | MW565217 |
| USA/CA-CZB-14768/2020 | EPI_ISL_739272 | MW565216 |
| USA/CA-CZB-14769/2020 | EPI_ISL_738844 | MW565215 |
| USA/CA-CZB-14770/2020 | EPI_ISL_738742 | MW565214 |
| USA/CA-CZB-14771/2020 | EPI_ISL_738852 | MW565213 |
| USA/CA-CZB-14772/2020 | EPI_ISL_739411 | MW565212 |
| USA/CA-CZB-14773/2020 | EPI_ISL_738827 | MW565211 |
| USA/CA-CZB-14774/2020 | EPI_ISL_738600 | MW565210 |
| USA/CA-CZB-14775/2020 | EPI_ISL_739527 | MW565209 |
| USA/CA-CZB-14777/2020 | EPI_ISL_739330 | MW565208 |
| USA/CA-CZB-14778/2020 | EPI_ISL_738912 | MW565207 |
| USA/CA-CZB-14779/2020 | EPI_ISL_738795 | MW565206 |
| USA/CA-CZB-14780/2020 | EPI_ISL_739367 | MW565205 |
| USA/CA-CZB-14781/2020 | EPI_ISL_738759 | MW565204 |
| USA/CA-CZB-14782/2020 | EPI_ISL_738985 | MW565203 |
| USA/CA-CZB-14783/2020 | EPI_ISL_738779 | MW565202 |
| USA/CA-CZB-14784/2020 | EPI_ISL_738810 | MW565201 |

| | | |
|---|---|---|
| USA/CA-CZB-14785/2020 | EPI_ISL_738913 | MW565200 |
| USA/CA-CZB-14786/2020 | EPI_ISL_739553 | MW565199 |
| USA/CA-CZB-14788/2020 | EPI_ISL_739194 | MW565198 |
| USA/CA-CZB-14789/2020 | EPI_ISL_739621 | MW565197 |
| USA/CA-CZB-14790/2020 | EPI_ISL_739525 | MW565196 |
| USA/CA-CZB-14791/2020 | EPI_ISL_738923 | MW565195 |
| USA/CA-CZB-14792/2020 | EPI_ISL_739032 | MW565194 |
| USA/CA-CZB-14793/2020 | EPI_ISL_739447 | MW565193 |
| USA/CA-CZB-14795/2020 | EPI_ISL_739318 | MW565192 |
| USA/CA-CZB-14796/2020 | EPI_ISL_739542 | MW565191 |
| USA/CA-CZB-14797/2020 | EPI_ISL_739389 | MW565190 |
| USA/CA-CZB-14798/2020 | EPI_ISL_2658662 | NA |
| USA/CA-CZB-14799/2020 | EPI_ISL_739210 | MW565189 |
| USA/CA-CZB-14801/2020 | EPI_ISL_739043 | MW565188 |
| USA/CA-CZB-14802/2020 | EPI_ISL_738806 | MW565187 |
| USA/CA-CZB-14803/2020 | EPI_ISL_739112 | MW565186 |
| USA/CA-CZB-14804/2020 | EPI_ISL_739611 | MW565185 |
| USA/CA-CZB-14805/2020 | EPI_ISL_738849 | MW565184 |
| USA/CA-CZB-14806/2020 | EPI_ISL_739436 | MW565183 |
| USA/CA-CZB-14807/2020 | EPI_ISL_738902 | MZ842389 |
| USA/CA-CZB-14808/2020 | EPI_ISL_739287 | MW565182 |
| USA/CA-CZB-14809/2020 | EPI_ISL_739313 | MW565181 |
| USA/CA-CZB-14810/2020 | EPI_ISL_739240 | MW565180 |
| USA/CA-CZB-14812/2020 | EPI_ISL_739000 | MW565179 |
| USA/CA-CZB-14813/2020 | EPI_ISL_739228 | MW565178 |
| USA/CA-CZB-14814/2020 | EPI_ISL_739531 | MW565177 |
| USA/CA-CZB-14815/2020 | EPI_ISL_738917 | MW565176 |
| USA/CA-CZB-14816/2020 | EPI_ISL_739170 | MW565175 |
| USA/CA-CZB-14817/2020 | EPI_ISL_739001 | MW565174 |

| | | |
|---|---|---|
| USA/CA-CZB-14818/2020 | EPI_ISL_739114 | MW565173 |
| USA/CA-CZB-14819/2020 | EPI_ISL_739007 | MW565172 |
| USA/CA-CZB-14820/2020 | EPI_ISL_739242 | MW565171 |
| USA/CA-CZB-14821/2020 | EPI_ISL_739651 | MW565170 |
| USA/CA-CZB-14822/2020 | EPI_ISL_738504 | MW565169 |
| USA/CA-CZB-14823/2020 | EPI_ISL_738875 | MW565168 |
| USA/CA-CZB-14824/2020 | EPI_ISL_738980 | MW565167 |
| USA/CA-CZB-14825/2020 | EPI_ISL_738865 | MW565166 |
| USA/CA-CZB-14826/2020 | EPI_ISL_739405 | MW565165 |
| USA/CA-CZB-14827/2020 | EPI_ISL_739403 | MW565164 |
| USA/CA-CZB-14828/2020 | EPI_ISL_738938 | MW565163 |
| USA/CA-CZB-14829/2020 | EPI_ISL_738807 | MW565162 |
| USA/CA-CZB-14830/2020 | EPI_ISL_739475 | MW565161 |
| USA/CA-CZB-14831/2020 | EPI_ISL_739417 | MW565160 |
| USA/CA-CZB-14832/2020 | EPI_ISL_739123 | MW565159 |
| USA/CA-CZB-14833/2020 | EPI_ISL_738801 | MW565158 |
| USA/CA-CZB-14834/2020 | EPI_ISL_739274 | MW565157 |
| USA/CA-CZB-14835/2020 | EPI_ISL_738563 | MW565156 |
| USA/CA-CZB-14836/2020 | EPI_ISL_738969 | MZ842388 |
| USA/CA-CZB-14837/2020 | EPI_ISL_739461 | MW565155 |
| USA/CA-CZB-14838/2020 | EPI_ISL_739425 | MW565154 |
| USA/CA-CZB-14839/2020 | EPI_ISL_738651 | MW565153 |
| USA/CA-CZB-14840/2020 | EPI_ISL_738539 | MW565152 |
| USA/CA-CZB-14841/2020 | EPI_ISL_739299 | MW565151 |
| USA/CA-CZB-14842/2020 | EPI_ISL_739183 | MW565150 |
| USA/CA-CZB-14844/2020 | EPI_ISL_739585 | MW565149 |
| USA/CA-CZB-14845/2020 | EPI_ISL_738649 | MW565148 |
| USA/CA-CZB-14846/2020 | EPI_ISL_739106 | MW565147 |
| USA/CA-CZB-14847/2020 | EPI_ISL_738929 | MW565146 |

| | | |
|---|---|---|
| USA/CA-CZB-14848/2020 | EPI_ISL_738647 | MW565145 |
| USA/CA-CZB-14849/2020 | EPI_ISL_738999 | MW565144 |
| USA/CA-CZB-14850/2020 | EPI_ISL_738854 | MZ842387 |
| USA/CA-CZB-14851/2020 | EPI_ISL_739452 | MW565143 |
| USA/CA-CZB-14852/2020 | EPI_ISL_739418 | MW565142 |
| USA/CA-CZB-14853/2020 | EPI_ISL_739155 | MW565141 |
| USA/CA-CZB-14854/2020 | EPI_ISL_739126 | MW565140 |
| USA/CA-CZB-14855/2020 | EPI_ISL_738958 | MW565139 |
| USA/CA-CZB-14856/2020 | EPI_ISL_739499 | MW565138 |
| USA/CA-CZB-14857/2020 | EPI_ISL_738809 | MW565137 |
| USA/CA-CZB-14858/2020 | EPI_ISL_738505 | MW565136 |
| USA/CA-CZB-14859/2020 | EPI_ISL_739250 | MW565135 |
| USA/CA-CZB-14860/2020 | EPI_ISL_739608 | MZ842386 |
| USA/CA-CZB-14861/2020 | EPI_ISL_739159 | MW565134 |
| USA/CA-CZB-14862/2020 | EPI_ISL_739198 | MW565133 |
| USA/CA-CZB-14864/2020 | EPI_ISL_739235 | MW565132 |
| USA/CA-CZB-14865/2020 | EPI_ISL_739131 | MW565131 |
| USA/CA-CZB-14866/2020 | EPI_ISL_739269 | MW565130 |
| USA/CA-CZB-1488/2020 | EPI_ISL_468438 | MT628172 |
| USA/CA-CZB-1489/2020 | EPI_ISL_468439 | NA |
| USA/CA-CZB-1490/2020 | EPI_ISL_468440 | MT628173 |
| USA/CA-CZB-1491/2020 | EPI_ISL_468441 | NA |
| USA/CA-CZB-1492/2020 | EPI_ISL_468442 | NA |
| USA/CA-CZB-1493/2020 | EPI_ISL_468443 | NA |
| USA/CA-CZB-1494/2020 | EPI_ISL_468444 | NA |
| USA/CA-CZB-1495/2020 | EPI_ISL_468445 | NA |
| USA/CA-CZB-1497/2020 | EPI_ISL_468446 | MT628174 |
| USA/CA-CZB-1498/2020 | EPI_ISL_468447 | MT628175 |
| USA/CA-CZB-1499/2020 | EPI_ISL_468448 | MT628176 |

| | | |
|---|---|---|
| USA/CA-CZB-1502/2020 | EPI_ISL_468449 | MT628177 |
| USA/CA-CZB-1503/2020 | EPI_ISL_468450 | MT628178 |
| USA/CA-CZB-1504/2020 | EPI_ISL_468451 | MT628179 |
| USA/CA-CZB-1505/2020 | EPI_ISL_468452 | MT628180 |
| USA/CA-CZB-1506/2020 | EPI_ISL_468453 | MT628181 |
| USA/CA-CZB-1507/2020 | EPI_ISL_468454 | MT628182 |
| USA/CA-CZB-1508/2020 | EPI_ISL_468455 | MT628183 |
| USA/CA-CZB-1509/2020 | EPI_ISL_468456 | MT628184 |
| USA/CA-CZB-1510/2020 | EPI_ISL_468457 | MT628185 |
| USA/CA-CZB-1511/2020 | EPI_ISL_468458 | MT628186 |
| USA/CA-CZB-1512/2020 | EPI_ISL_468459 | MT628187 |
| USA/CA-CZB-1513/2020 | EPI_ISL_468460 | MT628188 |
| USA/CA-CZB-1514/2020 | EPI_ISL_468461 | MT628189 |
| USA/CA-CZB-17317/2020 | EPI_ISL_955496 | MW702500 |
| USA/CA-CZB-17319/2020 | EPI_ISL_955497 | MW702499 |
| USA/CA-CZB-17320/2020 | EPI_ISL_1027535 | MW703127 |
| USA/CA-CZB-17321/2020 | EPI_ISL_955498 | MW702498 |
| USA/CA-CZB-17322/2020 | EPI_ISL_955499 | MW702497 |
| USA/CA-CZB-17323/2020 | EPI_ISL_955500 | MW702496 |
| USA/CA-CZB-17324/2020 | EPI_ISL_955501 | MW702495 |
| USA/CA-CZB-17325/2020 | EPI_ISL_955502 | MW702494 |
| USA/CA-CZB-17326/2020 | EPI_ISL_955503 | MW702493 |
| USA/CA-CZB-17327/2020 | EPI_ISL_955504 | MW702492 |
| USA/CA-CZB-17329/2020 | EPI_ISL_1027536 | MW703126 |
| USA/CA-CZB-17331/2020 | EPI_ISL_955505 | MW702491 |
| USA/CA-CZB-17332/2020 | EPI_ISL_955506 | MW702490 |
| USA/CA-CZB-17333/2020 | EPI_ISL_955507 | MW702489 |
| USA/CA-CZB-17334/2020 | EPI_ISL_955508 | MW702488 |
| USA/CA-CZB-17335/2020 | EPI_ISL_955509 | MW702487 |

| | | |
|---|---|---|
| USA/CA-CZB-17336/2020 | EPI_ISL_955510 | MW702486 |
| USA/CA-CZB-17337/2020 | EPI_ISL_955511 | MW702485 |
| USA/CA-CZB-17338/2020 | EPI_ISL_955512 | MW702484 |
| USA/CA-CZB-17339/2020 | EPI_ISL_955513 | MW702483 |
| USA/CA-CZB-17340/2020 | EPI_ISL_955514 | MW702482 |
| USA/CA-CZB-17341/2020 | EPI_ISL_1027537 | MW703125 |
| USA/CA-CZB-17342/2020 | EPI_ISL_955515 | MW702481 |
| USA/CA-CZB-17343/2020 | EPI_ISL_955516 | MW702480 |
| USA/CA-CZB-17344/2020 | EPI_ISL_955517 | MW702479 |
| USA/CA-CZB-17345/2020 | EPI_ISL_955518 | MW702478 |
| USA/CA-CZB-17346/2020 | EPI_ISL_955519 | MW702477 |
| USA/CA-CZB-17347/2020 | EPI_ISL_955520 | MW702476 |
| USA/CA-CZB-17348/2020 | EPI_ISL_1027538 | MW703124 |
| USA/CA-CZB-17349/2020 | EPI_ISL_955521 | MW702475 |
| USA/CA-CZB-17350/2020 | EPI_ISL_955522 | MW702474 |
| USA/CA-CZB-17351/2020 | EPI_ISL_955523 | MW702473 |
| USA/CA-CZB-17352/2020 | EPI_ISL_955524 | MW702472 |
| USA/CA-CZB-17353/2020 | EPI_ISL_955525 | MW702471 |
| USA/CA-CZB-17354/2020 | EPI_ISL_2658668 | NA |
| USA/CA-CZB-17355/2020 | EPI_ISL_955526 | MW702470 |
| USA/CA-CZB-17356/2020 | EPI_ISL_955527 | MW702469 |
| USA/CA-CZB-17357/2020 | EPI_ISL_955528 | MW702468 |
| USA/CA-CZB-17358/2020 | EPI_ISL_955529 | MW702467 |
| USA/CA-CZB-17359/2020 | EPI_ISL_955530 | MW702466 |
| USA/CA-CZB-17360/2020 | EPI_ISL_955531 | MW702465 |
| USA/CA-CZB-17361/2020 | EPI_ISL_955532 | MW702464 |
| USA/CA-CZB-17362/2020 | EPI_ISL_955533 | MW702463 |
| USA/CA-CZB-17363/2020 | EPI_ISL_955534 | MW702462 |
| USA/CA-CZB-17364/2020 | EPI_ISL_955535 | MW702461 |

| | | |
|---|---|---|
| USA/CA-CZB-17365/2020 | EPI_ISL_955536 | MW702460 |
| USA/CA-CZB-17366/2020 | EPI_ISL_955537 | MW702459 |
| USA/CA-CZB-17367/2020 | EPI_ISL_955538 | MW702458 |
| USA/CA-CZB-17368/2020 | EPI_ISL_955539 | MW702457 |
| USA/CA-CZB-17369/2020 | EPI_ISL_955540 | MW702456 |
| USA/CA-CZB-17370/2020 | EPI_ISL_955541 | MW702455 |
| USA/CA-CZB-17371/2020 | EPI_ISL_2659259 | MW702454 |
| USA/CA-CZB-17372/2020 | EPI_ISL_955542 | MW702453 |
| USA/CA-CZB-17373/2020 | EPI_ISL_2659308 | MW702452 |
| USA/CA-CZB-17374/2020 | EPI_ISL_955543 | MW702451 |
| USA/CA-CZB-17375/2020 | EPI_ISL_955544 | MW702450 |
| USA/CA-CZB-17376/2020 | EPI_ISL_955545 | MW702449 |
| USA/CA-CZB-17377/2020 | EPI_ISL_955546 | MW702448 |
| USA/CA-CZB-17378/2020 | EPI_ISL_955547 | MW702447 |
| USA/CA-CZB-17379/2020 | EPI_ISL_955548 | MW702446 |
| USA/CA-CZB-17380/2020 | EPI_ISL_1027540 | MW703123 |
| USA/CA-CZB-17381/2020 | EPI_ISL_955549 | MW702445 |
| USA/CA-CZB-17382/2020 | EPI_ISL_955550 | MW702444 |
| USA/CA-CZB-17383/2020 | EPI_ISL_955551 | MW702443 |
| USA/CA-CZB-17384/2020 | EPI_ISL_955552 | MW702442 |
| USA/CA-CZB-17385/2020 | EPI_ISL_955553 | MW702441 |
| USA/CA-CZB-17386/2020 | EPI_ISL_955554 | MW702440 |
| USA/CA-CZB-17387/2020 | EPI_ISL_955555 | MW702439 |
| USA/CA-CZB-17388/2020 | EPI_ISL_1027539 | MW703122 |
| USA/CA-CZB-17389/2020 | EPI_ISL_955556 | MW702438 |
| USA/CA-CZB-17390/2020 | EPI_ISL_955557 | MW702437 |
| USA/CA-CZB-17393/2020 | EPI_ISL_1027541 | MW703121 |
| USA/CA-CZB-17394/2020 | EPI_ISL_955558 | NA |
| USA/CA-CZB-17395/2020 | EPI_ISL_955559 | MW702436 |

| | | |
|---|---|---|
| USA/CA-CZB-17397/2020 | EPI_ISL_955560 | MW702435 |
| USA/CA-CZB-19111/2020 | EPI_ISL_979710 | MZ842239 |
| USA/CA-CZB-19112/2020 | EPI_ISL_979711 | MW701299 |
| USA/CA-CZB-19113/2020 | EPI_ISL_979712 | MW701298 |
| USA/CA-CZB-19114/2020 | EPI_ISL_979713 | MW701297 |
| USA/CA-CZB-19115/2020 | EPI_ISL_979714 | MW701296 |
| USA/CA-CZB-19116/2020 | EPI_ISL_979715 | MW701295 |
| USA/CA-CZB-19117/2020 | EPI_ISL_979716 | MW701294 |
| USA/CA-CZB-19118/2020 | EPI_ISL_979717 | MW701293 |
| USA/CA-CZB-19119/2020 | EPI_ISL_979718 | MW701292 |
| USA/CA-CZB-19120/2020 | EPI_ISL_2659251 | MW701291 |
| USA/CA-CZB-19121/2020 | EPI_ISL_979719 | MW701290 |
| USA/CA-CZB-19123/2020 | EPI_ISL_2659384 | MW701289 |
| USA/CA-CZB-19124/2020 | EPI_ISL_979720 | MW701288 |
| USA/CA-CZB-19125/2020 | EPI_ISL_979721 | MW701287 |
| USA/CA-CZB-19126/2020 | EPI_ISL_979722 | MW701286 |
| USA/CA-CZB-19127/2020 | EPI_ISL_979723 | MW701285 |
| USA/CA-CZB-19128/2020 | EPI_ISL_979724 | MW701284 |
| USA/CA-CZB-19129/2020 | EPI_ISL_979725 | MW701283 |
| USA/CA-CZB-19132/2020 | EPI_ISL_979726 | MW701282 |
| USA/CA-CZB-19133/2020 | EPI_ISL_979727 | MW701281 |
| USA/CA-CZB-19134/2020 | EPI_ISL_979728 | MW701280 |
| USA/CA-CZB-19135/2020 | EPI_ISL_979729 | MW701279 |
| USA/CA-CZB-19136/2020 | EPI_ISL_979730 | MW701278 |
| USA/CA-CZB-19137/2020 | EPI_ISL_979731 | MW701277 |
| USA/CA-CZB-19138/2020 | EPI_ISL_979732 | MW701276 |
| USA/CA-CZB-19139/2020 | EPI_ISL_979733 | MW701275 |
| USA/CA-CZB-19140/2020 | EPI_ISL_979734 | MW701274 |
| USA/CA-CZB-19141/2020 | EPI_ISL_979735 | MW701273 |

| | | |
|---|---|---|
| USA/CA-CZB-19142/2020 | EPI_ISL_979736 | MW701272 |
| USA/CA-CZB-19143/2020 | EPI_ISL_979737 | MW701271 |
| USA/CA-CZB-19144/2020 | EPI_ISL_979738 | MW701270 |
| USA/CA-CZB-19145/2020 | EPI_ISL_979739 | MW701269 |
| USA/CA-CZB-19146/2020 | EPI_ISL_979740 | MW701268 |
| USA/CA-CZB-19147/2020 | EPI_ISL_979741 | MW701267 |
| USA/CA-CZB-19148/2020 | EPI_ISL_979742 | MW701266 |
| USA/CA-CZB-19149/2020 | EPI_ISL_979743 | MW701265 |
| USA/CA-CZB-19150/2020 | EPI_ISL_979744 | MW701264 |
| USA/CA-CZB-19151/2020 | EPI_ISL_979745 | MW701263 |
| USA/CA-CZB-19152/2020 | EPI_ISL_979746 | MW701262 |
| USA/CA-CZB-19153/2020 | EPI_ISL_979747 | MW701261 |
| USA/CA-CZB-19154/2020 | EPI_ISL_979748 | MW701260 |
| USA/CA-CZB-19155/2020 | EPI_ISL_979749 | MW701259 |
| USA/CA-CZB-19156/2020 | EPI_ISL_979750 | MW701258 |
| USA/CA-CZB-19157/2020 | EPI_ISL_979751 | MW701257 |
| USA/CA-CZB-19158/2020 | EPI_ISL_979752 | MW701256 |
| USA/CA-CZB-19159/2020 | EPI_ISL_979753 | MW701255 |
| USA/CA-CZB-19160/2020 | EPI_ISL_979754 | MW701254 |
| USA/CA-CZB-19161/2020 | EPI_ISL_979755 | MW701253 |
| USA/CA-CZB-19162/2020 | EPI_ISL_979756 | MW701252 |
| USA/CA-CZB-19163/2020 | EPI_ISL_979757 | MW701251 |
| USA/CA-CZB-19164/2020 | EPI_ISL_979758 | MW701250 |
| USA/CA-CZB-19165/2020 | EPI_ISL_979759 | MW701249 |
| USA/CA-CZB-19166/2020 | EPI_ISL_979760 | MW701248 |
| USA/CA-CZB-19167/2020 | EPI_ISL_979761 | MW701247 |
| USA/CA-CZB-19168/2020 | EPI_ISL_979762 | MW701246 |
| USA/CA-CZB-19169/2020 | EPI_ISL_979763 | MW701245 |
| USA/CA-CZB-19170/2020 | EPI_ISL_979764 | MW701244 |

| | | |
|---|---|---|
| USA/CA-CZB-19171/2020 | EPI_ISL_979765 | MW701243 |
| USA/CA-CZB-19172/2020 | EPI_ISL_979766 | MW701242 |
| USA/CA-CZB-19173/2020 | EPI_ISL_979767 | MW701241 |
| USA/CA-CZB-19174/2020 | EPI_ISL_979768 | MW701240 |
| USA/CA-CZB-19175/2020 | EPI_ISL_979769 | MW701239 |
| USA/CA-CZB-19176/2020 | EPI_ISL_979770 | MW701238 |
| USA/CA-CZB-19177/2020 | EPI_ISL_979771 | MW701237 |
| USA/CA-CZB-19178/2020 | EPI_ISL_979772 | MW701236 |
| USA/CA-CZB-19179/2020 | EPI_ISL_979773 | MW701235 |
| USA/CA-CZB-19180/2020 | EPI_ISL_979774 | MW701234 |
| USA/CA-CZB-19181/2020 | EPI_ISL_979775 | MW701233 |
| USA/CA-CZB-19182/2020 | EPI_ISL_979776 | MW701232 |
| USA/CA-CZB-19183/2020 | EPI_ISL_979777 | MW701231 |
| USA/CA-CZB-19184/2020 | EPI_ISL_979778 | MW701230 |
| USA/CA-CZB-19185/2020 | EPI_ISL_979779 | MW701229 |
| USA/CA-CZB-19186/2020 | EPI_ISL_979780 | MW701228 |
| USA/CA-CZB-19187/2020 | EPI_ISL_979781 | MW701227 |
| USA/CA-CZB-19188/2020 | EPI_ISL_979782 | MW701226 |
| USA/CA-CZB-19189/2020 | EPI_ISL_979783 | MW701225 |
| USA/CA-CZB-19190/2020 | EPI_ISL_979784 | MW701224 |
| USA/CA-CZB-19191/2020 | EPI_ISL_2758426 | MZ842238 |
| USA/CA-CZB-19192/2020 | EPI_ISL_979785 | MW701223 |
| USA/CA-CZB-19193/2020 | EPI_ISL_979786 | MW701222 |
| USA/CA-CZB-19194/2020 | EPI_ISL_979787 | MW701221 |
| USA/CA-CZB-19195/2020 | EPI_ISL_979788 | MW701220 |
| USA/CA-CZB-19196/2020 | EPI_ISL_979789 | MW701219 |
| USA/CA-CZB-19197/2020 | EPI_ISL_979790 | MW701218 |
| USA/CA-CZB-19199/2020 | EPI_ISL_979791 | MW701217 |
| USA/CA-CZB-19200/2020 | EPI_ISL_979792 | MW701216 |

| | | |
|---|---|---|
| USA/CA-CZB-19201/2020 | EPI_ISL_979793 | MW701215 |
| USA/CA-CZB-19202/2020 | EPI_ISL_979794 | MW701214 |
| USA/CA-CZB-19203/2020 | EPI_ISL_979795 | MW701213 |
| USA/CA-CZB-19204/2020 | EPI_ISL_979796 | MW701212 |
| USA/CA-CZB-19205/2020 | EPI_ISL_979797 | MW701211 |
| USA/CA-CZB-19207/2020 | EPI_ISL_979162 | MW700768 |
| USA/CA-CZB-19208/2020 | EPI_ISL_979163 | MW700767 |
| USA/CA-CZB-19209/2020 | EPI_ISL_979164 | MW700766 |
| USA/CA-CZB-19210/2020 | EPI_ISL_979165 | MW700765 |
| USA/CA-CZB-19211/2020 | EPI_ISL_979166 | MW700764 |
| USA/CA-CZB-19212/2020 | EPI_ISL_979167 | MW700763 |
| USA/CA-CZB-19214/2020 | EPI_ISL_979168 | MW700762 |
| USA/CA-CZB-19215/2020 | EPI_ISL_979169 | MW700761 |
| USA/CA-CZB-19216/2020 | EPI_ISL_1030499 | MW702982 |
| USA/CA-CZB-19217/2020 | EPI_ISL_979170 | MW700760 |
| USA/CA-CZB-19218/2020 | EPI_ISL_979171 | MW700759 |
| USA/CA-CZB-19219/2020 | EPI_ISL_979172 | NA |
| USA/CA-CZB-19220/2020 | EPI_ISL_979173 | MW700758 |
| USA/CA-CZB-19221/2020 | EPI_ISL_979174 | MW700757 |
| USA/CA-CZB-19222/2020 | EPI_ISL_979175 | MW700756 |
| USA/CA-CZB-19223/2020 | EPI_ISL_979176 | MW700755 |
| USA/CA-CZB-19224/2020 | EPI_ISL_979177 | MW700754 |
| USA/CA-CZB-19225/2020 | EPI_ISL_979178 | MW700753 |
| USA/CA-CZB-19227/2020 | EPI_ISL_979179 | MW700752 |
| USA/CA-CZB-19228/2020 | EPI_ISL_979180 | MW700751 |
| USA/CA-CZB-19229/2020 | EPI_ISL_979181 | MW700750 |
| USA/CA-CZB-19230/2020 | EPI_ISL_979182 | MW700749 |
| USA/CA-CZB-19232/2020 | EPI_ISL_979183 | MW700748 |
| USA/CA-CZB-19233/2020 | EPI_ISL_979184 | MW700747 |

| | | |
|---|---|---|
| USA/CA-CZB-19235/2020 | EPI_ISL_979185 | MW700746 |
| USA/CA-CZB-19236/2020 | EPI_ISL_979186 | MW700745 |
| USA/CA-CZB-19237/2020 | EPI_ISL_979187 | MW700744 |
| USA/CA-CZB-19238/2020 | EPI_ISL_979188 | MW700743 |
| USA/CA-CZB-19239/2020 | EPI_ISL_979189 | MW700742 |
| USA/CA-CZB-19240/2020 | EPI_ISL_979190 | MW700741 |
| USA/CA-CZB-19241/2020 | EPI_ISL_979191 | MW700740 |
| USA/CA-CZB-19242/2020 | EPI_ISL_979192 | MW700739 |
| USA/CA-CZB-19243/2020 | EPI_ISL_979193 | MW700738 |
| USA/CA-CZB-19244/2020 | EPI_ISL_979194 | MW700737 |
| USA/CA-CZB-19246/2020 | EPI_ISL_979195 | MW700736 |
| USA/CA-CZB-19247/2020 | EPI_ISL_979196 | MW700735 |
| USA/CA-CZB-19248/2020 | EPI_ISL_979197 | MW700734 |
| USA/CA-CZB-19249/2020 | EPI_ISL_979198 | MW700733 |
| USA/CA-CZB-19250/2020 | EPI_ISL_979199 | MW700732 |
| USA/CA-CZB-19251/2020 | EPI_ISL_979200 | MW700731 |
| USA/CA-CZB-19252/2020 | EPI_ISL_979201 | MW700730 |
| USA/CA-CZB-19253/2020 | EPI_ISL_979202 | MW700729 |
| USA/CA-CZB-19254/2020 | EPI_ISL_979203 | MW700728 |
| USA/CA-CZB-19255/2020 | EPI_ISL_979204 | MW700727 |
| USA/CA-CZB-19256/2020 | EPI_ISL_979205 | MW700726 |
| USA/CA-CZB-19257/2020 | EPI_ISL_979206 | MW700725 |
| USA/CA-CZB-19258/2020 | EPI_ISL_979207 | MZ842237 |
| USA/CA-CZB-19259/2020 | EPI_ISL_979208 | MW700724 |
| USA/CA-CZB-19261/2020 | EPI_ISL_979209 | MW700723 |
| USA/CA-CZB-19262/2020 | EPI_ISL_979210 | MW700722 |
| USA/CA-CZB-19263/2020 | EPI_ISL_2658604 | NA |
| USA/CA-CZB-19265/2020 | EPI_ISL_979211 | MW700721 |
| USA/CA-CZB-19266/2020 | EPI_ISL_979212 | MW700720 |

| | | |
|---|---|---|
| USA/CA-CZB-19267/2020 | EPI_ISL_979213 | MW700719 |
| USA/CA-CZB-19268/2020 | EPI_ISL_979214 | MW700718 |
| USA/CA-CZB-19269/2020 | EPI_ISL_979215 | MW700717 |
| USA/CA-CZB-19271/2020 | EPI_ISL_979216 | MW700716 |
| USA/CA-CZB-19272/2020 | EPI_ISL_979217 | MW700715 |
| USA/CA-CZB-19273/2020 | EPI_ISL_979218 | MW700714 |
| USA/CA-CZB-19274/2020 | EPI_ISL_979219 | MW700713 |
| USA/CA-CZB-19275/2020 | EPI_ISL_979220 | MW700712 |
| USA/CA-CZB-19276/2020 | EPI_ISL_979221 | MW700711 |
| USA/CA-CZB-19277/2020 | EPI_ISL_979222 | MW700710 |
| USA/CA-CZB-19278/2020 | EPI_ISL_979223 | MW700709 |
| USA/CA-CZB-19279/2020 | EPI_ISL_979224 | MW700708 |
| USA/CA-CZB-19280/2020 | EPI_ISL_979225 | MW700707 |
| USA/CA-CZB-19281/2020 | EPI_ISL_979226 | MW700706 |
| USA/CA-CZB-19282/2020 | EPI_ISL_979227 | MW700705 |
| USA/CA-CZB-19283/2020 | EPI_ISL_979228 | MW700704 |
| USA/CA-CZB-19284/2020 | EPI_ISL_979229 | MW700703 |
| USA/CA-CZB-19285/2020 | EPI_ISL_979230 | MW700702 |
| USA/CA-CZB-19286/2020 | EPI_ISL_979231 | MW700701 |
| USA/CA-CZB-19287/2020 | EPI_ISL_979232 | MZ842236 |
| USA/CA-CZB-19288/2020 | EPI_ISL_979233 | MZ842235 |
| USA/CA-CZB-19289/2020 | EPI_ISL_979234 | MW700700 |
| USA/CA-CZB-19290/2020 | EPI_ISL_979235 | MW700699 |
| USA/CA-CZB-19291/2020 | EPI_ISL_979236 | MW700698 |
| USA/CA-CZB-19292/2020 | EPI_ISL_979237 | MW700697 |
| USA/CA-CZB-19293/2020 | EPI_ISL_979238 | MW700696 |
| USA/CA-CZB-19294/2020 | EPI_ISL_979239 | MW700695 |
| USA/CA-CZB-19295/2020 | EPI_ISL_979240 | MW700694 |
| USA/CA-CZB-19296/2020 | EPI_ISL_979241 | MW700693 |

| | | |
|---|---|---|
| USA/CA-CZB-19297/2020 | EPI_ISL_979242 | MW700692 |
| USA/CA-CZB-19298/2020 | EPI_ISL_979243 | MW700691 |
| USA/CA-CZB-19299/2020 | EPI_ISL_979244 | MW700690 |
| USA/CA-CZB-19301/2020 | EPI_ISL_979245 | MW700689 |
| USA/CA-CZB-19302/2020 | EPI_ISL_955693 | MW700467 |
| USA/CA-CZB-19303/2020 | EPI_ISL_2758427 | MZ842234 |
| USA/CA-CZB-19304/2020 | EPI_ISL_955694 | MW700466 |
| USA/CA-CZB-19305/2020 | EPI_ISL_955695 | MW700465 |
| USA/CA-CZB-19306/2020 | EPI_ISL_955696 | MW700464 |
| USA/CA-CZB-19307/2020 | EPI_ISL_955697 | MW700463 |
| USA/CA-CZB-19308/2020 | EPI_ISL_955698 | MW700462 |
| USA/CA-CZB-19309/2020 | EPI_ISL_955699 | MW700461 |
| USA/CA-CZB-19310/2020 | EPI_ISL_955700 | MW700460 |
| USA/CA-CZB-19311/2020 | EPI_ISL_955701 | MW700459 |
| USA/CA-CZB-19312/2020 | EPI_ISL_955702 | MW700458 |
| USA/CA-CZB-19313/2020 | EPI_ISL_955703 | MW700457 |
| USA/CA-CZB-19314/2020 | EPI_ISL_955704 | MW700456 |
| USA/CA-CZB-19315/2020 | EPI_ISL_955705 | MW700455 |
| USA/CA-CZB-19316/2020 | EPI_ISL_955706 | MW700454 |
| USA/CA-CZB-19317/2020 | EPI_ISL_955707 | MW700453 |
| USA/CA-CZB-19318/2020 | EPI_ISL_955708 | MW700452 |
| USA/CA-CZB-19319/2020 | EPI_ISL_955709 | MW700451 |
| USA/CA-CZB-19320/2020 | EPI_ISL_955710 | MW700450 |
| USA/CA-CZB-19321/2020 | EPI_ISL_955711 | MW700449 |
| USA/CA-CZB-19322/2020 | EPI_ISL_955712 | MW700448 |
| USA/CA-CZB-19323/2020 | EPI_ISL_955713 | MW700447 |
| USA/CA-CZB-19324/2020 | EPI_ISL_955714 | MW700446 |
| USA/CA-CZB-19325/2020 | EPI_ISL_955715 | MW700445 |
| USA/CA-CZB-19326/2020 | EPI_ISL_955716 | MW700444 |

| | | |
|---|---|---|
| USA/CA-CZB-19327/2020 | EPI_ISL_955717 | MW700443 |
| USA/CA-CZB-19328/2020 | EPI_ISL_955718 | MW700442 |
| USA/CA-CZB-19329/2020 | EPI_ISL_955719 | MW700441 |
| USA/CA-CZB-19330/2020 | EPI_ISL_955720 | MW700440 |
| USA/CA-CZB-19331/2020 | EPI_ISL_955721 | MW700439 |
| USA/CA-CZB-19332/2020 | EPI_ISL_955722 | MW700438 |
| USA/CA-CZB-19333/2020 | EPI_ISL_955723 | MW700437 |
| USA/CA-CZB-19334/2020 | EPI_ISL_1027561 | MW703037 |
| USA/CA-CZB-19335/2020 | EPI_ISL_2659213 | MW700436 |
| USA/CA-CZB-19336/2020 | EPI_ISL_955724 | MW700435 |
| USA/CA-CZB-19338/2020 | EPI_ISL_955725 | MW700434 |
| USA/CA-CZB-19339/2020 | EPI_ISL_955726 | MW700433 |
| USA/CA-CZB-19340/2020 | EPI_ISL_955727 | MW700432 |
| USA/CA-CZB-19341/2020 | EPI_ISL_2758428 | MZ842233 |
| USA/CA-CZB-19342/2020 | EPI_ISL_955728 | MW700431 |
| USA/CA-CZB-19343/2020 | EPI_ISL_955729 | MW700430 |
| USA/CA-CZB-19344/2020 | EPI_ISL_955730 | MW700429 |
| USA/CA-CZB-19345/2020 | EPI_ISL_2659635 | MW700428 |
| USA/CA-CZB-19346/2020 | EPI_ISL_955731 | MW700427 |
| USA/CA-CZB-19347/2020 | EPI_ISL_955732 | MW700426 |
| USA/CA-CZB-19348/2020 | EPI_ISL_955733 | MW700425 |
| USA/CA-CZB-19349/2020 | EPI_ISL_955734 | MW700424 |
| USA/CA-CZB-19350/2020 | EPI_ISL_955735 | MW700423 |
| USA/CA-CZB-19351/2020 | EPI_ISL_955736 | MW700422 |
| USA/CA-CZB-19352/2020 | EPI_ISL_955737 | MW700421 |
| USA/CA-CZB-19353/2020 | EPI_ISL_955738 | MW700420 |
| USA/CA-CZB-19354/2020 | EPI_ISL_955739 | MW700419 |
| USA/CA-CZB-19355/2020 | EPI_ISL_955740 | MW700418 |
| USA/CA-CZB-19356/2020 | EPI_ISL_955741 | MW700417 |

| | | |
|---|---|---|
| USA/CA-CZB-19357/2020 | EPI_ISL_955742 | MW700416 |
| USA/CA-CZB-19358/2020 | EPI_ISL_955743 | MW700415 |
| USA/CA-CZB-19359/2020 | EPI_ISL_955744 | MW700414 |
| USA/CA-CZB-19360/2020 | EPI_ISL_955745 | MW700413 |
| USA/CA-CZB-19361/2020 | EPI_ISL_955746 | MW700412 |
| USA/CA-CZB-19362/2020 | EPI_ISL_955747 | MW700411 |
| USA/CA-CZB-19363/2020 | EPI_ISL_955748 | MW700410 |
| USA/CA-CZB-19364/2020 | EPI_ISL_955749 | MW700409 |
| USA/CA-CZB-19365/2020 | EPI_ISL_955750 | MZ842232 |
| USA/CA-CZB-19366/2020 | EPI_ISL_955751 | MW700408 |
| USA/CA-CZB-19367/2020 | EPI_ISL_955752 | MW700407 |
| USA/CA-CZB-19368/2020 | EPI_ISL_955753 | MW700406 |
| USA/CA-CZB-19369/2020 | EPI_ISL_2658710 | NA |
| USA/CA-CZB-19371/2020 | EPI_ISL_955754 | MW700405 |
| USA/CA-CZB-19372/2020 | EPI_ISL_955755 | MW700404 |
| USA/CA-CZB-19373/2020 | EPI_ISL_955756 | MW700403 |
| USA/CA-CZB-19374/2020 | EPI_ISL_955757 | MW700402 |
| USA/CA-CZB-19375/2020 | EPI_ISL_955758 | MW700401 |
| USA/CA-CZB-19376/2020 | EPI_ISL_955759 | MW700400 |
| USA/CA-CZB-19377/2020 | EPI_ISL_955760 | MW700399 |
| USA/CA-CZB-19378/2020 | EPI_ISL_955761 | MW700398 |
| USA/CA-CZB-19379/2020 | EPI_ISL_955762 | MW700397 |
| USA/CA-CZB-19380/2020 | EPI_ISL_955763 | MW700396 |
| USA/CA-CZB-19381/2021 | EPI_ISL_955764 | MW700395 |
| USA/CA-CZB-19382/2020 | EPI_ISL_955765 | MW700394 |
| USA/CA-CZB-19383/2020 | EPI_ISL_955766 | MW700393 |
| USA/CA-CZB-19384/2021 | EPI_ISL_955767 | MW700392 |
| USA/CA-CZB-19385/2021 | EPI_ISL_955768 | MW700391 |
| USA/CA-CZB-19387/2021 | EPI_ISL_955769 | MW700390 |

| | | |
|---|---|---|
| USA/CA-CZB-19388/2021 | EPI_ISL_955770 | MW700389 |
| USA/CA-CZB-19389/2021 | EPI_ISL_955771 | MW700388 |
| USA/CA-CZB-19390/2021 | EPI_ISL_955772 | MW700387 |
| USA/CA-CZB-19391/2021 | EPI_ISL_955773 | MW700386 |
| USA/CA-CZB-19392/2021 | EPI_ISL_955774 | MW700385 |
| USA/CA-CZB-19393/2021 | EPI_ISL_1027476 | MW703036 |
| USA/CA-CZB-19394/2021 | EPI_ISL_955775 | MW700384 |
| USA/CA-CZB-19395/2021 | EPI_ISL_955776 | MW700383 |
| USA/CA-CZB-19396/2021 | EPI_ISL_1027517 | MW703035 |
| USA/CA-CZB-19397/2021 | EPI_ISL_955777 | MW700382 |
| USA/CA-CZB-2033/2020 | EPI_ISL_1924619 | MZ842848 |
| USA/CA-CZB-2034/2020 | EPI_ISL_486280 | MT750454 |
| USA/CA-CZB-2035/2020 | EPI_ISL_486281 | MZ842847 |
| USA/CA-CZB-2036/2020 | EPI_ISL_486282 | MT750455 |
| USA/CA-CZB-2037/2020 | EPI_ISL_486283 | MT750456 |
| USA/CA-CZB-2039/2020 | EPI_ISL_486284 | MZ842846 |
| USA/CA-CZB-2041/2020 | EPI_ISL_486285 | MT750457 |
| USA/CA-CZB-2042/2020 | EPI_ISL_486286 | MT750458 |
| USA/CA-CZB-23111/2021 | EPI_ISL_1185409 | MW739359 |
| USA/CA-CZB-23112/2021 | EPI_ISL_1185355 | MW739358 |
| USA/CA-CZB-23113/2021 | EPI_ISL_1185438 | MW739357 |
| USA/CA-CZB-23115/2021 | EPI_ISL_1185428 | MW739356 |
| USA/CA-CZB-23116/2021 | EPI_ISL_1185294 | MW739355 |
| USA/CA-CZB-23117/2021 | EPI_ISL_1185302 | MW739354 |
| USA/CA-CZB-23118/2021 | EPI_ISL_1185528 | MW739353 |
| USA/CA-CZB-23119/2021 | EPI_ISL_1185359 | MW739352 |
| USA/CA-CZB-23120/2021 | EPI_ISL_1185489 | MW739351 |
| USA/CA-CZB-23122/2021 | EPI_ISL_1185361 | MW739350 |
| USA/CA-CZB-23123/2021 | EPI_ISL_1185408 | MW739349 |

| | | |
|---|---|---|
| USA/CA-CZB-23124/2021 | EPI_ISL_1185497 | MW739348 |
| USA/CA-CZB-23125/2021 | EPI_ISL_1185448 | MW739347 |
| USA/CA-CZB-23126/2021 | EPI_ISL_1185505 | MW739346 |
| USA/CA-CZB-23127/2021 | EPI_ISL_1185382 | MW739345 |
| USA/CA-CZB-23128/2021 | EPI_ISL_1185385 | MW739344 |
| USA/CA-CZB-23129/2021 | EPI_ISL_1185319 | MW739343 |
| USA/CA-CZB-23130/2021 | EPI_ISL_1185318 | MW739342 |
| USA/CA-CZB-23131/2021 | EPI_ISL_1185325 | MW739341 |
| USA/CA-CZB-23132/2021 | EPI_ISL_1185395 | MW739340 |
| USA/CA-CZB-23133/2021 | EPI_ISL_1185304 | MW739339 |
| USA/CA-CZB-23134/2021 | EPI_ISL_1185345 | MW739338 |
| USA/CA-CZB-23135/2021 | EPI_ISL_1185455 | MW739337 |
| USA/CA-CZB-23136/2021 | EPI_ISL_1185352 | MW739336 |
| USA/CA-CZB-23137/2021 | EPI_ISL_1185399 | MW739335 |
| USA/CA-CZB-23138/2021 | EPI_ISL_1185298 | MW739334 |
| USA/CA-CZB-23139/2021 | EPI_ISL_2659557 | MW739333 |
| USA/CA-CZB-23140/2021 | EPI_ISL_1185460 | MZ842204 |
| USA/CA-CZB-23141/2021 | EPI_ISL_1185369 | MW739332 |
| USA/CA-CZB-23142/2021 | EPI_ISL_1185488 | MW739331 |
| USA/CA-CZB-23143/2021 | EPI_ISL_1185326 | MW739330 |
| USA/CA-CZB-23144/2021 | EPI_ISL_1185468 | MW739329 |
| USA/CA-CZB-23145/2021 | EPI_ISL_2659338 | MW739328 |
| USA/CA-CZB-23146/2021 | EPI_ISL_1185312 | MW739327 |
| USA/CA-CZB-23147/2021 | EPI_ISL_1185471 | MW739326 |
| USA/CA-CZB-23148/2021 | EPI_ISL_1185309 | MW739325 |
| USA/CA-CZB-23149/2021 | EPI_ISL_1185300 | MW739324 |
| USA/CA-CZB-23150/2021 | EPI_ISL_1185301 | MW739323 |
| USA/CA-CZB-23151/2021 | EPI_ISL_1185292 | MW739322 |
| USA/CA-CZB-23152/2021 | EPI_ISL_2659380 | MW739321 |

| | | |
|---|---|---|
| USA/CA-CZB-23153/2021 | EPI_ISL_2659359 | MW739320 |
| USA/CA-CZB-23155/2021 | EPI_ISL_2659316 | MW739319 |
| USA/CA-CZB-23156/2021 | EPI_ISL_1185474 | MW739318 |
| USA/CA-CZB-23157/2021 | EPI_ISL_1185365 | MW739317 |
| USA/CA-CZB-23158/2021 | EPI_ISL_1185454 | MW739316 |
| USA/CA-CZB-23159/2021 | EPI_ISL_1185394 | MW739315 |
| USA/CA-CZB-23161/2021 | EPI_ISL_1185509 | MW739314 |
| USA/CA-CZB-23162/2021 | EPI_ISL_1185436 | MW739313 |
| USA/CA-CZB-23163/2021 | EPI_ISL_1185407 | MW739312 |
| USA/CA-CZB-23164/2021 | EPI_ISL_1185308 | MW739311 |
| USA/CA-CZB-23165/2021 | EPI_ISL_1185346 | MW739310 |
| USA/CA-CZB-23166/2021 | EPI_ISL_1185507 | MW739309 |
| USA/CA-CZB-23167/2021 | EPI_ISL_1185417 | MW739308 |
| USA/CA-CZB-23168/2021 | EPI_ISL_2659319 | MW739307 |
| USA/CA-CZB-23169/2021 | EPI_ISL_2659336 | MW739306 |
| USA/CA-CZB-23170/2021 | EPI_ISL_2659609 | MW739305 |
| USA/CA-CZB-23171/2021 | EPI_ISL_1185526 | MW739304 |
| USA/CA-CZB-23172/2021 | EPI_ISL_1185360 | MW739303 |
| USA/CA-CZB-23173/2021 | EPI_ISL_1185287 | MW739302 |
| USA/CA-CZB-23174/2021 | EPI_ISL_1185476 | MW739301 |
| USA/CA-CZB-23175/2021 | EPI_ISL_1185440 | MW739300 |
| USA/CA-CZB-23177/2021 | EPI_ISL_1185329 | MW739299 |
| USA/CA-CZB-23178/2021 | EPI_ISL_1185424 | MW739298 |
| USA/CA-CZB-23179/2021 | EPI_ISL_1185291 | MW739297 |
| USA/CA-CZB-23180/2021 | EPI_ISL_1185479 | MW739296 |
| USA/CA-CZB-23181/2021 | EPI_ISL_1185481 | MW739295 |
| USA/CA-CZB-23182/2021 | EPI_ISL_1185469 | MW739294 |
| USA/CA-CZB-23183/2021 | EPI_ISL_1185323 | MW739293 |
| USA/CA-CZB-23184/2021 | EPI_ISL_1185353 | MW739292 |

| | | |
|---|---|---|
| USA/CA-CZB-23185/2021 | EPI_ISL_1185519 | MW739291 |
| USA/CA-CZB-23186/2021 | EPI_ISL_1185521 | MW739290 |
| USA/CA-CZB-23187/2021 | EPI_ISL_1185328 | MW739289 |
| USA/CA-CZB-23188/2021 | EPI_ISL_1185504 | MW739288 |
| USA/CA-CZB-23189/2021 | EPI_ISL_1185413 | MW739287 |
| USA/CA-CZB-23190/2021 | EPI_ISL_1185410 | MW739286 |
| USA/CA-CZB-23191/2021 | EPI_ISL_2659421 | MW739285 |
| USA/CA-CZB-23192/2021 | EPI_ISL_1185340 | MW739284 |
| USA/CA-CZB-23193/2021 | EPI_ISL_1185498 | MW739283 |
| USA/CA-CZB-23194/2021 | EPI_ISL_1185425 | MW739282 |
| USA/CA-CZB-23195/2021 | EPI_ISL_1185324 | MZ842203 |
| USA/CA-CZB-23196/2021 | EPI_ISL_1185516 | MW739281 |
| USA/CA-CZB-23198/2021 | EPI_ISL_1185432 | MW739280 |
| USA/CA-CZB-23199/2021 | EPI_ISL_1185518 | MW739279 |
| USA/CA-CZB-23200/2021 | EPI_ISL_1185486 | MW739278 |
| USA/CA-CZB-23201/2021 | EPI_ISL_1185487 | MW739277 |
| USA/CA-CZB-23202/2021 | EPI_ISL_1185463 | MW739276 |
| USA/CA-CZB-23203/2021 | EPI_ISL_1185306 | MW739275 |
| USA/CA-CZB-23204/2021 | EPI_ISL_1185439 | MW739274 |
| USA/CA-CZB-23205/2021 | EPI_ISL_2659265 | MW739273 |
| USA/CA-CZB-23206/2021 | EPI_ISL_1185412 | MW739272 |
| USA/CA-CZB-2419/2020 | EPI_ISL_513840 | MZ842842 |
| USA/CA-CZB-2420/2020 | EPI_ISL_513841 | MW035992 |
| USA/CA-CZB-2421/2020 | EPI_ISL_513842 | MW035999 |
| USA/CA-CZB-2422/2020 | EPI_ISL_513843 | MW036022 |
| USA/CA-CZB-2423/2020 | EPI_ISL_513844 | MW036010 |
| USA/CA-CZB-2424/2020 | EPI_ISL_513845 | MW035997 |
| USA/CA-CZB-2425/2020 | EPI_ISL_513846 | MW036024 |
| USA/CA-CZB-2426/2020 | EPI_ISL_513847 | MW036047 |

| | | |
|---|---|---|
| USA/CA-CZB-2429/2020 | EPI_ISL_513848 | MW036038 |
| USA/CA-CZB-2430/2020 | EPI_ISL_513849 | MW035991 |
| USA/CA-CZB-2431/2020 | EPI_ISL_513850 | MW036084 |
| USA/CA-CZB-2433/2020 | EPI_ISL_513851 | NA |
| USA/CA-CZB-2436/2020 | EPI_ISL_513852 | MW036056 |
| USA/CA-CZB-2443/2020 | EPI_ISL_513853 | MW036090 |
| USA/CA-CZB-2445/2020 | EPI_ISL_513854 | MW036003 |
| USA/CA-CZB-2446/2020 | EPI_ISL_513855 | MW036073 |
| USA/CA-CZB-25659/2021 | EPI_ISL_1234789 | MW739803 |
| USA/CA-CZB-25660/2021 | EPI_ISL_1234794 | MW739802 |
| USA/CA-CZB-25661/2021 | EPI_ISL_1234799 | MW739801 |
| USA/CA-CZB-25662/2021 | EPI_ISL_1235011 | MW739800 |
| USA/CA-CZB-25663/2021 | EPI_ISL_1234833 | MW739799 |
| USA/CA-CZB-25664/2021 | EPI_ISL_1234871 | MW739798 |
| USA/CA-CZB-25665/2021 | EPI_ISL_2658600 | NA |
| USA/CA-CZB-25666/2021 | EPI_ISL_1234807 | MW739797 |
| USA/CA-CZB-25667/2021 | EPI_ISL_1234930 | MW739796 |
| USA/CA-CZB-25668/2021 | EPI_ISL_1234826 | MW739795 |
| USA/CA-CZB-25669/2021 | EPI_ISL_1234831 | MW739794 |
| USA/CA-CZB-25670/2021 | EPI_ISL_1234793 | MW739793 |
| USA/CA-CZB-25671/2021 | EPI_ISL_1235012 | MW739792 |
| USA/CA-CZB-25672/2021 | EPI_ISL_1235023 | MW739791 |
| USA/CA-CZB-25674/2021 | EPI_ISL_1234955 | MW739790 |
| USA/CA-CZB-25675/2021 | EPI_ISL_1234949 | MW739789 |
| USA/CA-CZB-25676/2021 | EPI_ISL_1234836 | MW739788 |
| USA/CA-CZB-25678/2021 | EPI_ISL_1235002 | MW739787 |
| USA/CA-CZB-25679/2021 | EPI_ISL_1234908 | MW739786 |
| USA/CA-CZB-25680/2021 | EPI_ISL_1235030 | MW739785 |
| USA/CA-CZB-25681/2021 | EPI_ISL_1235046 | MW739784 |

| | | |
|---|---|---|
| USA/CA-CZB-25682/2021 | EPI_ISL_1234788 | MW739783 |
| USA/CA-CZB-25683/2021 | EPI_ISL_1235004 | MW739782 |
| USA/CA-CZB-25684/2021 | EPI_ISL_2658582 | NA |
| USA/CA-CZB-25685/2021 | EPI_ISL_1234896 | MW739781 |
| USA/CA-CZB-25686/2021 | EPI_ISL_1234898 | MW739780 |
| USA/CA-CZB-25687/2021 | EPI_ISL_1234922 | MW739779 |
| USA/CA-CZB-25688/2021 | EPI_ISL_1234852 | MW739778 |
| USA/CA-CZB-25689/2021 | EPI_ISL_1235039 | MW739777 |
| USA/CA-CZB-25690/2021 | EPI_ISL_1234927 | MW739776 |
| USA/CA-CZB-25691/2021 | EPI_ISL_1235037 | MW739775 |
| USA/CA-CZB-25692/2021 | EPI_ISL_1235003 | MW739774 |
| USA/CA-CZB-25693/2021 | EPI_ISL_1234859 | MW739773 |
| USA/CA-CZB-25694/2021 | EPI_ISL_1234998 | MW739772 |
| USA/CA-CZB-25695/2021 | EPI_ISL_1234948 | MW739771 |
| USA/CA-CZB-25696/2021 | EPI_ISL_1234889 | MW739770 |
| USA/CA-CZB-25697/2021 | EPI_ISL_1234823 | MW739769 |
| USA/CA-CZB-25698/2021 | EPI_ISL_1234981 | MW739768 |
| USA/CA-CZB-25699/2021 | EPI_ISL_1234960 | MW739767 |
| USA/CA-CZB-25701/2021 | EPI_ISL_1234882 | MW739766 |
| USA/CA-CZB-25702/2021 | EPI_ISL_1234868 | MW739765 |
| USA/CA-CZB-25703/2021 | EPI_ISL_1234989 | MW739764 |
| USA/CA-CZB-25704/2021 | EPI_ISL_1235001 | MW739763 |
| USA/CA-CZB-25707/2021 | EPI_ISL_1235005 | MW739762 |
| USA/CA-CZB-25708/2021 | EPI_ISL_2758452 | MZ842102 |
| USA/CA-CZB-25709/2021 | EPI_ISL_1235040 | MW739761 |
| USA/CA-CZB-25710/2021 | EPI_ISL_1234824 | MW739760 |
| USA/CA-CZB-25711/2021 | EPI_ISL_1234861 | MW739759 |
| USA/CA-CZB-25712/2021 | EPI_ISL_1234971 | MW739758 |
| USA/CA-CZB-25713/2021 | EPI_ISL_1234801 | MW739757 |

| | | |
|---|---|---|
| USA/CA-CZB-25714/2021 | EPI_ISL_1234903 | MW739756 |
| USA/CA-CZB-25715/2021 | EPI_ISL_1234910 | MW739755 |
| USA/CA-CZB-25716/2021 | EPI_ISL_1234853 | MW739754 |
| USA/CA-CZB-25718/2021 | EPI_ISL_1234811 | MW739753 |
| USA/CA-CZB-25719/2021 | EPI_ISL_1234934 | MW739752 |
| USA/CA-CZB-25720/2021 | EPI_ISL_1234912 | MW739751 |
| USA/CA-CZB-25721/2021 | EPI_ISL_1234814 | MW739750 |
| USA/CA-CZB-25722/2021 | EPI_ISL_1235033 | MW739749 |
| USA/CA-CZB-25723/2021 | EPI_ISL_1234911 | MW739748 |
| USA/CA-CZB-25724/2021 | EPI_ISL_1234803 | MW739747 |
| USA/CA-CZB-25725/2021 | EPI_ISL_1234874 | MW739746 |
| USA/CA-CZB-25726/2021 | EPI_ISL_2658646 | NA |
| USA/CA-CZB-25727/2021 | EPI_ISL_1234936 | MW739745 |
| USA/CA-CZB-25728/2021 | EPI_ISL_1234916 | MW739744 |
| USA/CA-CZB-25729/2021 | EPI_ISL_1234822 | MW739743 |
| USA/CA-CZB-25730/2021 | EPI_ISL_1234980 | MW739742 |
| USA/CA-CZB-25731/2021 | EPI_ISL_1234805 | MW739741 |
| USA/CA-CZB-25732/2021 | EPI_ISL_1234786 | MW739740 |
| USA/CA-CZB-25733/2021 | EPI_ISL_1234817 | MW739739 |
| USA/CA-CZB-25734/2021 | EPI_ISL_1234906 | MW739738 |
| USA/CA-CZB-25735/2021 | EPI_ISL_1234816 | MW739737 |
| USA/CA-CZB-25736/2021 | EPI_ISL_1234863 | MW739736 |
| USA/CA-CZB-25737/2021 | EPI_ISL_1234890 | MW739735 |
| USA/CA-CZB-25738/2021 | EPI_ISL_1235045 | MW739734 |
| USA/CA-CZB-25739/2021 | EPI_ISL_1234821 | MW739733 |
| USA/CA-CZB-25740/2021 | EPI_ISL_1234931 | MW739732 |
| USA/CA-CZB-25741/2021 | EPI_ISL_1235053 | MW739731 |
| USA/CA-CZB-25742/2021 | EPI_ISL_1234928 | MW739730 |
| USA/CA-CZB-25743/2021 | EPI_ISL_1234888 | MW739729 |

| | | |
|---|---|---|
| USA/CA-CZB-25744/2021 | EPI_ISL_1234893 | MW739728 |
| USA/CA-CZB-25745/2021 | EPI_ISL_1234984 | MW739727 |
| USA/CA-CZB-25746/2021 | EPI_ISL_1234914 | MW739726 |
| USA/CA-CZB-25747/2021 | EPI_ISL_1234905 | MW739725 |
| USA/CA-CZB-25748/2021 | EPI_ISL_2758453 | MZ842101 |
| USA/CA-CZB-25749/2021 | EPI_ISL_1234792 | MW739724 |
| USA/CA-CZB-25750/2021 | EPI_ISL_1234966 | MW739723 |
| USA/CA-CZB-25751/2021 | EPI_ISL_1234963 | MW739722 |
| USA/CA-CZB-25752/2021 | EPI_ISL_1234856 | MW739721 |
| USA/CA-CZB-25753/2021 | EPI_ISL_2659310 | MZ842100 |
| USA/CA-CZB-25754/2021 | EPI_ISL_1234974 | MW739720 |
